# Cell-free DNA ultra-low-pass whole genome sequencing to distinguish malignant peripheral nerve sheath tumor (MPNST) from its benign precursor lesion: a cross-sectional study

**DOI:** 10.1101/2021.04.22.21255769

**Authors:** Jeffrey J. Szymanski, R. Taylor Sundby, Paul A. Jones, Divya Srihari, Noah Earland, Peter K. Harris, Wenjia Feng, Faridi Qaium, Haiyan Lei, David Roberts, Michele Landeau, Jamie Bell, Yi Huang, Leah Hoffman, Melissa Spencer, Matthew B. Spraker, Li Ding, Brigitte C. Widemann, Jack F. Shern, Angela C. Hirbe, Aadel A. Chaudhuri

## Abstract

**Background:** The leading cause of mortality for patients with the Neurofibromatosis type 1 (NF1) cancer predisposition syndrome is development of Malignant Peripheral Nerve Sheath Tumor (MPNST), an aggressive soft tissue sarcoma. In the setting of NF1, this cancer type frequently arises from within its common and benign precursor, plexiform neurofibroma (PN). Transformation from PN to MPNST is challenging to diagnose due to difficulties in distinguishing cross-sectional imaging results and intralesional heterogeneity resulting in biopsy sampling errors.

**Methods and Findings:** This multi-institutional study from the National Cancer Institute and Washington University in St. Louis used fragment size analysis and ultra-low-pass whole genome sequencing (ULP-WGS) of plasma cell-free DNA (cfDNA) to distinguish between MPNST and PN in patients with NF1. Following in-silico enrichment for short cfDNA fragments and copy number analysis to estimate the fraction of plasma cfDNA originating from tumor (tumor fraction), we developed a noninvasive classifier which differentiates MPNST from PN with 86% pre-treatment accuracy (91% specificity, 75% sensitivity), and 89% accuracy on serial analysis (91% specificity, 83% sensitivity). Healthy controls without NF1 (subjects = 16, plasma samples = 16), PN (subjects = 23, plasma samples = 23), and MPNST (subjects = 14, plasma samples = 46) cohorts showed significant differences in tumor fraction in plasma (*P* = 0.001 as well as cfDNA fragment length (*P* < 0.001) with MPNST samples harboring shorter fragments and being enriched for tumor-derived cfDNA relative to PN and healthy controls. Mutational analysis demonstrated focal NF1 copy number loss in PN and MPNST patient plasma but not in healthy controls. Greater genomic instability including alterations associated with malignant transformation (focal copy number gains in chromosome arms 1q, 7p, 8q, 9q, and 17q; focal copy number losses in *SUZ12, SMARCA2, CDKN2A/B*, and chromosome arms 6p and 9p) was more prominently observed in MPNST plasma. Furthermore, the sum of longest tumor diameters (SLD) visualized by cross-sectional imaging correlated significantly with paired tumor fractions in plasma from MPNST patients (*r* = 0.39, *P* = 0.024). On serial analysis, tumor fraction levels in plasma dynamically correlated with treatment response to therapy and minimal residual disease (MRD) detection before relapse. Study limitations include a modest MPNST sample size despite accrual from two major referral centers for this rare malignancy, and lack of uniform treatment and imaging protocols representing a real-world cohort.

**Conclusions:** Tumor fraction levels derived from cfDNA fragment size and copy number alteration analysis of plasma cfDNA using ULP-WGS significantly correlated with MPNST tumor burden, accurately distinguished MPNST from its benign PN precursor, and dynamically correlated with treatment response. In the future, our findings could form the basis for improved early cancer detection and monitoring in high-risk cancer-predisposed populations.

**Author summary:** *Why was this study done?:*

➢ Neurofibromatosis 1 (NF1) is the most common inherited cancer predisposition syndrome.
➢ The leading cause of mortality in NF1 is malignant peripheral nerve sheath tumor (MPNST), an aggressive soft tissue sarcoma that arises from a benign plexiform neurofibroma (PN) precursor lesion.
➢ Transformation from PN to MPNST is challenging to detect by imaging (due to difficulty in distinguishing PN from MPNST radiologically) or by biopsy (due to intralesional heterogeneity), which often delays the diagnosis of MPNST and results in a worsened prognosis.

*What did the researchers do and find?:*

➢ We conducted a multi-institutional study involving two large NF1 referral centers, the National Cancer Institute and Washington University in St. Louis, involving 73 patients from whom plasma cell-free DNA (cfDNA) was analyzed using ultra-low-pass whole genome sequencing (ULP-WGS).
We found that cfDNA from patients with MPNST harbors a shorter fragmentation profile compared to patients with PN or healthy donors. Using sequencing reads from this fragmentation profile, we quantified genome-wide copy number alterations (CNAs) in cfDNA and used CNAs to estimate the fraction of plasma cfDNA originating from tumor.
➢ Tumor fraction in plasma cfDNA distinguished pre-treatment MPSNT from PN with 86% accuracy. Plasma cfDNA from MPNST and PN patients harbored focal copy number loss of NF1, not found in healthy donors. Strikingly, MPNST patient cfDNA also had significantly greater tumor genomic instability compared to PN, with copy number alterations in key genomic loci previously observed in MPNST tissue (i.e., gain of chromosome arm 8q and loss of 9p), which enabled sensitive and specific liquid biopsy discrimination of MPNST from PN.
➢ Plasma-derived tumor fraction correlated with tumor size from imaging in MPNST patients, and serial cfDNA analysis demonstrated the potential for noninvasive detection of minimal residual disease, treatment response assessment, and the potential for even greater assay sensitivity.

*What do these findings mean?:*

➢ Our findings suggest that cfDNA fragment analysis followed by ULP-WGS has the potential to be developed as a biomarker for treatment response, and as a screening assay for early detection of MPNST.
➢ This study provides, to our knowledge, the first evidence for the ability of liquid biopsy to distinguish between benign and malignant tumors in a heritable cancer predisposition syndrome.

## Introduction

Neurofibromatosis type 1 (NF1) is an autosomal dominant disorder affecting one in 3,000 individuals worldwide, and is caused by a heterozygous inactivating mutation in the tumor suppressor gene, *NF1*, located on chromosome 17q11.2 [1-3]. *NF1* encodes for the protein, neurofibromin 1, a negative regulator of the RAS signaling pathway. Thus, *NF1* loss-of-function mutations lead to hyperactivated RAS, whose downstream effects contribute to the elevated cancer risk in NF1 patients [4-6].

Approximately 50% of patients with NF1 develop histologically benign plexiform neurofibroma (PN) [1,7], in which Schwann cells acquire biallelic inactivation of the *NF1* gene [3,8]. Histologically, PNs are heterogeneous, consisting of primarily S100-positive Schwann cells (60–80%), as well as fibroblasts, endothelial cells, perineural cells, smooth muscle cells, mast cells, interspersed axons, and pericytes [2]. Imaging studies of PN mirror this heterogeneity, complicating the radiographic diagnosis of transformation to malignant peripheral nerve sheath tumor (MPNST), which occurs in 8-15% of patients with NF1 [1,9,10], as well as the accuracy of diagnostic tissue biopsy.

MPNST are aggressive cancers with a poor prognosis that frequently arise from within their benign PN precursors [9,11-13]. Due to rapid development of metastasis and resistance to both chemotherapy and radiotherapy, MPNST account for the majority of NF1-associated mortality [1,9] with a 5-year survival rate of only 20% [14]. Despite the high incidence and mortality of MPNST in the NF1 population, screening for malignant transformation and monitoring of MPNST is challenging. Clinical exam has poor sensitivity and may only signify MPNST when a PN lesion is showing sudden growth or causing severe pain [12,15]. Serial PN biopsies are impractical as 9-21% of NF1 patients will have multiple PN, with varying levels of malignant potential requiring surveillance [16-18]. Moreover, biopsies can yield false negative results due to geographic tumor heterogeneity resulting from MPNST arising from within heterogeneous PN precursor lesions [19]. Furthermore, standard cross-sectional imaging cannot distinguish MPNST from PN with adequate specificity [20,21]. Given the high prevalence of deadly MPNST in the context of a very common benign precursor lesion in a cancer-predisposed population, it is imperative that more reliable screening modalities be explored.

We and others have shown that other cancer types can be monitored through plasma cell-free DNA (cfDNA) analysis [22-25] and that malignancy can be associated with distinct cfDNA fragmentation profiles, typically characterized by shorter size [26-30]. We have also shown that sequenced MPNST tissue harbors broad chromosomal copy number alterations (CNAs) characteristic of increased genomic instability compared to PN, including in cases of MPNST transformation arising from within PN lesions [31,32]. Here, we hypothesize that this MPNST-intrinsic genomic instability is also detectable within plasma cfDNA, and can be used to noninvasively distinguish MPNST from its benign precursor lesion.

In the current multi-institutional cross-sectional study involving two large referral centers for NF1 patients, the Washington University School of Medicine and the National Cancer Institute, we aimed to develop a non-invasive liquid biopsy method for distinguishing MPNST from its benign PN precursor using cfDNA fragmentomics and ultra-low-pass whole genome sequencing (ULP-WGS).

## Patients and Methods

### Study design

This study used blood samples prospectively collected from NF1 patients with MPNST and PN tumors with the aim of distinguishing these different tumor types by plasma cell-free DNA analysis (**Figure 1**). Patients from the National Cancer Institute (NCI) and Washington University in St. Louis (WUSTL) with clinically and radiographically diagnosed PN or biopsy-proven MPNST were enrolled onto this multi-institutional cross-sectional study with written informed consent (NCI protocol NCT01109394, NIH Intramural IRB identifier 10C0086; NCI protocol NCT00924196, NIH Intramural IRB identifier 08C0079; WUSTL protocol NCT04354064, Washington University in St. Louis School of Medicine Human Research Protection Office IRB identifiers 201903142 and 201203042) between 2016 and 2020. NF1 status was determined clinically by consensus criteria [33]. Five subjects with MPNST who did not meet NF1 consensus criteria were also included in the analysis. A total of 14 MPNST and 23 PN patients were enrolled with peripheral blood collected at the time of enrollment (**Supplementary Tables 1-3**). MPNST patients had serial plasma samples collected for a total of 46 MPNST plasma samples (average 3, maximum 6 per subject, **Supplementary Table 1**). When available, tissue was also collected at a single timepoint (n = 4 subjects). When peripheral blood mononuclear cells (PBMCs) were isolated from whole blood, these were sequenced as germline DNA (n = 17 subjects). All patients underwent clinical management and follow-up by board-certified physicians per the standard-of-care. All samples were collected with informed consent for research and institutional review board approval in accordance with the Declaration of Helsinki. Protocols are available on ClinicalTrials.gov. A STROBE checklist was completed to assure an accurate and complete report of the study (see **Supplement**) [34].

**Figure 1:**
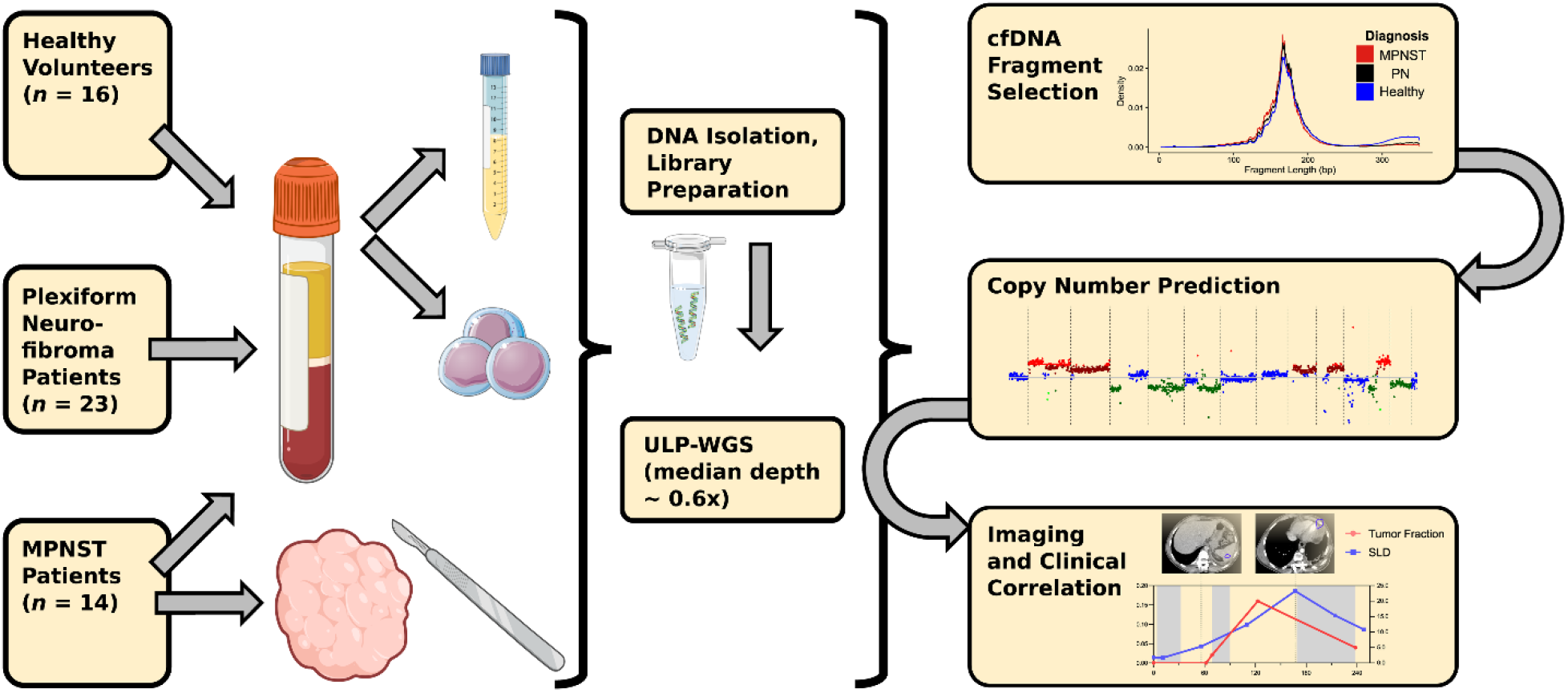
Study schema. Patients with imaging-and biopsy-proven MPNST and established PN along with healthy donors were enrolled onto this multi-institutional prospective cohort, with plasma collected for tumor fraction analysis at the time of study enrollment. Tumor fraction was assessed in all collected plasma samples by ULP-WGS followed by in-silico size selection for short cfDNA fragments, which was used to train a noninvasive MPNST vs. PN classifier. During subsequent treatment and follow-up, MPNST patients underwent further serial imaging (analyzed by RECIST) and plasma sample collection (analyzed by ULP-WGS and in-silico fragment size selection), with results correlated with each other and with clinical outcomes. cfDNA, cell-free DNA; MPNST, malignant peripheral nerve sheath tumor; PN, plexiform neurofibroma; RECIST, response evaluation criteria in solid tumors, version 1.1; ULP-WGS, ultra-low-pass whole genome sequencing.

### Healthy controls

After obtaining written consent, healthy donor blood samples were obtained at a single timepoint from appropriately consented donors at the NIH Department of Transfusion medicine (NIH protocol NCT00001846, NIH Intramural IRB identifier 99-CC-0168) and WUSTL Clinical Translational Research Unit (WUSTL protocol NCT04354064, Washington University in St. Louis School of Medicine Human Research Protection Office IRB identifiers 201903142 and 201203042) (**Supplementary Table 4**). Eligibility for healthy controls included age greater than 18 years old and no known history of neoplastic or hematological disorders. Protocols are available on ClinicalTrials.gov.

### Clinical specimens

After obtaining written informed consent for genomic analysis, serial peripheral blood samples were collected throughout the clinical course for consenting MPNST patients or at a single timepoint for PN patients and healthy controls. Treatment regimen for MPNST was determined by the primary treating clinicians, and included radiotherapy, surgery, and cytotoxic chemotherapy (**Supplementary Table 2**).

Venous blood samples (10-30 ml) were collected in EDTA (BD Biosciences, San Jose, CA) or Cell-Free DNA BCT (Streck Laboratories, La Vista, NE) tubes. EDTA tubes were processed within 4 hours of collection, while Cell-Free DNA BCT tubes were processed within 7 days of collection. Whole blood samples were centrifuged at room temperature (NCI: 1,900 x g for 10 minutes, WUSTL: 1,200 x g for 10 minutes). Isolated plasma was centrifuged a second time at room temperature (NCI: 15,000 x g for 10 minutes, WUSTL: 1,800 x g for 5 minutes) in low-bind Eppendorf tubes to remove residual cells. Purified plasma was frozen at −80 °C until cfDNA isolation.

### Plasma cell-free DNA isolation

Purified plasma was thawed at room temperature and cfDNA was extracted from 2-8 mL of plasma using the QIAamp Circulating Nucleic Acid kit (Qiagen, Hilden, Germany). Extracted DNA concentration was measured using the Qubit dsDNA High-Sensitivity assay (ThermoFisher, Waltham, MA) and cfDNA concentration and quality were assessed using a Bioanalyzer (Agilent Technologies, Santa Clara, CA) or Tapestation (Agilent Technologies, Santa Clara, CA). Isolated cfDNA was stored at –20°C until library preparation.

### Germline DNA isolation and processing

After centrifuging clinical venous blood samples and removing plasma supernatant per above, the red blood cells and buffy coat were resuspended in PBS for germline DNA extraction using the DNeasy Blood and Tissue kit (Qiagen, Hilden, Germany). For a subset of samples, germline DNA from peripheral blood mononuclear cells was collected in and extracted using PAXgene Blood DNA tubes and kit (PreAnalytix, Germantown, MD). DNA was stored at -20°C until further processing. Germline DNA was then fragmented using a LE220 focused ultrasonicator (Covaris, Woburn, MA) or a Q800R3 sonicator (Qsonica LLC, Newton, CT) according to the manufacturer’s instructions and previously published methods [35] to a target length of 200 bp. DNA lengths were assessed using a Bioanalyzer (Agilent Technologies, Santa Clara, CA).

### Tumor DNA isolation and processing

Tumor tissue was not procured for research unless clinically indicated and available following the standard clinical pathology workflow. When available, tumor tissue was snap-frozen and stored at −80°C or stored in formalin-fixed paraffin-embedding (FFPE). Nucleic acids were isolated from tumor FFPE samples using the manufacturer’s protocol with the AllPrep DNA/RNA FFPE kit (Qiagen, Hilden, Germany). DNA was extracted from snap-frozen tumor tissue samples using the DNeasy Blood and Tissue kit (Qiagen, Hilden, Germany). Extracted DNA was stored at -20°C until further processing. Tissue DNA was subsequently fragmented using a LE220 focused ultrasonicator (Covaris, Woburn, MA or Q800R3 sonicator (Qsonica LLC, Newton, CT) and analyzed using a Bioanalyzer (Agilent Technologies, Santa Clara, CA) as described above.

### DNA library construction and sequencing

Sequencing libraries were constructed from cfDNA (NCI 5-15 ng, WUSTL 10-60 ng) or germline/tumor DNA (NCI 100 ng, WUSTL 32 ng) using commercial kits per the manufacturers’ instructions: TruSeq Nano (Illumina, San Diego, CA) for NCI samples and Kapa HyperPrep (Roche, Basel, Switzerland) for WUSTL samples. Constructed libraries were balanced, pooled and sequenced using 150 bp paired-end reads on a NovaSeq (Illumina, San Diego, CA) or HiSeq 4000 (Illumina, San Diego, CA). Data was then quality-filtered and pooled for analysis.

### Copy number alteration and tumor fraction analysis

Sequencing data was demultiplexed and raw reads were quality-filtered using fastp v.0.2. Quality-filtered reads were then aligned to the hg19 human genome assembly using BWA v.0.7.17. Aligned reads were de-duplicated with Samtools v.1.7, then downsampled to 10 million read pairs (WGS coverage ∼0.6x), or separately for comparison purposes to 5 million read pairs (WGS coverage ∼0.3x). Genomic coverage was estimated using MosDepth [36]. To enrich for circulating tumor DNA (ctDNA) fragments, *in-silico* size selection was applied to all cfDNA samples [28]. Only quality-filtered reads between fragment lengths of 90 and 150 bp were considered for copy number and tumor fraction analysis for cfDNA samples, while such size selection was not performed for tumor and germline samples. GC-content and mappability bias correction, depth-based local copy number estimates, and copy number-based estimation of tumor fraction were then performed using the ichorCNA tool (Broad v.0.2.0) [37]. Briefly, reads were summed in non-overlapping windows of 10^6^ bases. Local read depth was corrected for GC bias and known regions of low mappability, and artifacts were removed by comparison to ichorCNA’s built-in healthy control reference. CNAs were predicted using recommended low tumor fraction parameters for cfDNA samples and default parameters for tumor and germline samples. X and Y chromosomes were not considered in copy number ratios. ichorCNA then used these binned, bias-corrected copy number values to model a two-component mixture of tumor-derived and non-tumor-derived fragments, from which it inferred the fraction of reads in each sample originating from tumor (tumor fraction) [37]. Visualization of genome-wide copy number alterations at specific loci (**Figure 3**) was generated from compiled log_2_ ratios of copy number for all study plasma specimens (*n* = 107 samples). Reads were classified as copy number gain if log_2_ of the copy number ratio was > 0.58 (log_2_ (3/2)) and loss if log_2_ of the copy number ratio was < –1.0 (log_2_ (1/2)). Large bin copy number alteration plots (**Supplementary Figure 1**) reflect copy number changes from baseline in the tumor-only fraction of each sample. Both copy number state and tumor fraction were determined by ichorCNA [37].

### Fragment size analysis

Following the sequencing quality-control, deduplication, alignment and downsampling steps described above, read-pair fragment sizes for cfDNA samples were calculated using deepTools bamPEFragmentSize [38]. The distribution of each sample’s fragment sizes was estimated by kernel density. Distributions were compared between the three clinical states (healthy control, PN, and MPNST), and between high and low tumor fraction samples by two-sided Kolmogorov-Smirnov testing.

### Comparisons of cfDNA tumor fraction to imaging

Patients with MPNST and PN were monitored by CT, MRI, and/or FDG-PET imaging at the treating institution at the managing clinicians’ discretion. For patients with MPNST, radiographic tumor burden was quantified by sum of the longest tumor diameters (SLD) per RECIST 1.1 criteria [39]. For comparison to serial timepoint cfDNA tumor fractions, each plasma sample was matched to the nearest SLD value at the primary institution within 30 days and without any interceding change of therapy. SLDs and plasma tumor fraction levels were then assessed using Pearson’s correlation coefficient. For comparisons of plasma tumor fraction to clinical status by RECIST, tumor fraction values were first normalized per patient to the lowest value detected on serial analysis, and then log_2_-transformed to generate the final plotted values in **Figure 5B**. Changes in clinical status were assessed and categorized as complete response, partial response, stable disease or progressive disease per RECIST 1.1 criteria [39]. RECIST 1.1 scoring was performed on serial imaging studies relative to a patient’s baseline scan.

### Power and statistical analyses

Previous tissue-based studies have shown that PN harbor few genome-wide CNAs [40,41] but acquire significant genomic instability during malignant transformation to MPNST [32,41,42]. Based on these known significant CNA differences between MPNST and PN tumors, we assumed a large effect size would also be evident comparing MPNST plasma tumor fraction to plasma from PN patients or healthy controls. Using Cohen’s *f* = 0.6 with an α = 0.05 and power = 0.80, we projected that the sample size needed to detect differences between these three categories would be *n* = 10 per group. Our category group sizes met or exceeded this estimate for all comparisons (**Supplementary Tables 1-4**).

When testing associations between plasma tumor fraction and clinical status (**Figure 4**), we limited MPNST plasma samples to those collected either prior to all treatments or after a washout period of least 21 days after completion of chemotherapy or radiation therapy (designated as pretreatment or baseline MPNST samples below). The distributions of plasma tumor fraction for each clinical status were compared by Kruskal-Wallis H test with pairwise comparisons by Dunn’s test. To further compare pretreatment MPNST to benign PN patients, we generated a receiver operating characteristic (ROC) curve of plasma tumor fraction. Tumor fraction values derived from ctDNA-enriched 90-150 bp fragments were compared to tumor fractions derived from all cfDNA fragment lengths. For ctDNA-enriched tumor fraction, an optimized cut-point was determined by Youden’s index (the point on the ROC curve that maximizes sensitivity + specificity – 1), and high and low-plasma tumor fraction groups by cut-point were compared to clinical status by Fisher’s exact test. A logistic regression was also performed for the MPNST versus PN groups using the glm function in *R*, evaluating the effects of age, sex and institution in addition to pretreatment plasma tumor fraction. Leave-one-out cross-validation was performed in *R* using the caret package. The reverse Kaplan-Meier method was used to estimate follow-up times. Statistical analyses were performed using *R* v.3.6.1 or Prism 9 (GraphPad Software).

## Results

### Overview and patient characteristics

The primary objective of this study was to noninvasively differentiate MPNST from benign PN by analyzing and quantifying genomic CNAs in blood plasma cfDNA (**Figure 1**). To quantify CNAs, we profiled 107 biospecimens including 85 plasma samples from 53 subjects by ULP-WGS downsampled to 10 million paired reads (∼0.6x) (**Figure 1; Supplementary Table 1**). Subject groups compared were MPNST and PN patients as well as healthy donor controls. Specimen types included blood plasma cell-free DNA, blood leukocyte germline DNA, and frozen tumor specimens (**Supplementary Table 1**). The median age was 36, 27 and 32.5 for MPNST patients, PN patients and healthy donors, respectively (**Supplementary Tables 2-4**). Exclusion criteria included diagnosis of a non-MPNST malignancy. No PN patients developed any clinical or radiographic evidence of MPNST transformation during a median study follow-up time of nearly 2 years (median 690 days; interquartile range of 531-1,140 days; **Supplementary Table 3**). For two subjects, sar015 and sar037, large PN resection was performed for lesion-related morbidity with final pathology confirming the PN diagnosis. Among patients with MPNST, 86.7% received chemotherapy, 35.7% received radiation therapy, and 42.9% underwent surgical resection (**Supplementary Table 2**).

### Genome-wide CNAs from tumor are detected in plasma

Eighty-six percent of MPNST and plexiform neurofibroma patients enrolled onto our study met the NIH criteria for NF1 diagnosis. There was no difference in tumor fraction between the MPNST patients who met NIH criteria and those who did not (*P* = 0.93 by Wilcoxon rank-sum test). Genomic copy number analysis of plasma cell-free DNA revealed that focal somatic CNAs that have previously been associated with PN tumor progression in NF1 patients [32] were prominently observed in MPNST patients and were occasionally found in PN patients, but absent in healthy controls (**Figures 2 and 3**). For example, focal loss of *CDKN2A/B, MTAP, SMARCA2*, and *SUZ12*, alterations shown to be associated with malignant transformation of PN [32,43,44], was observed in plasma from MPNST patients. *CDKN2B and SUZ12* losses were also found in two PN patients while *SMARCA2* appeared to be copy number neutral in plasma across the full PN cohort. Loss of *SUZ12* correlated with *NF1* loss, consistent with both genes’ location in the 17.q11.2 genomic locus [32,45]. Additionally, we observed broader copy number gains in chromosome arms 1q, 7p, 8q, 9q and 17q as well as broad losses in arms 6p and 9p only in MPNST patient plasma, again consistent with previous findings from NF1 MPNST tumors [32,42,46] (**Figure 3**). Finally, while many types of *NF1* gene activation can underlie the NF1 disease process, we observed evidence of one of these, *NF1* copy number loss, only within MPNST and PN patients, but not in healthy donor controls.

**Figure 2.**
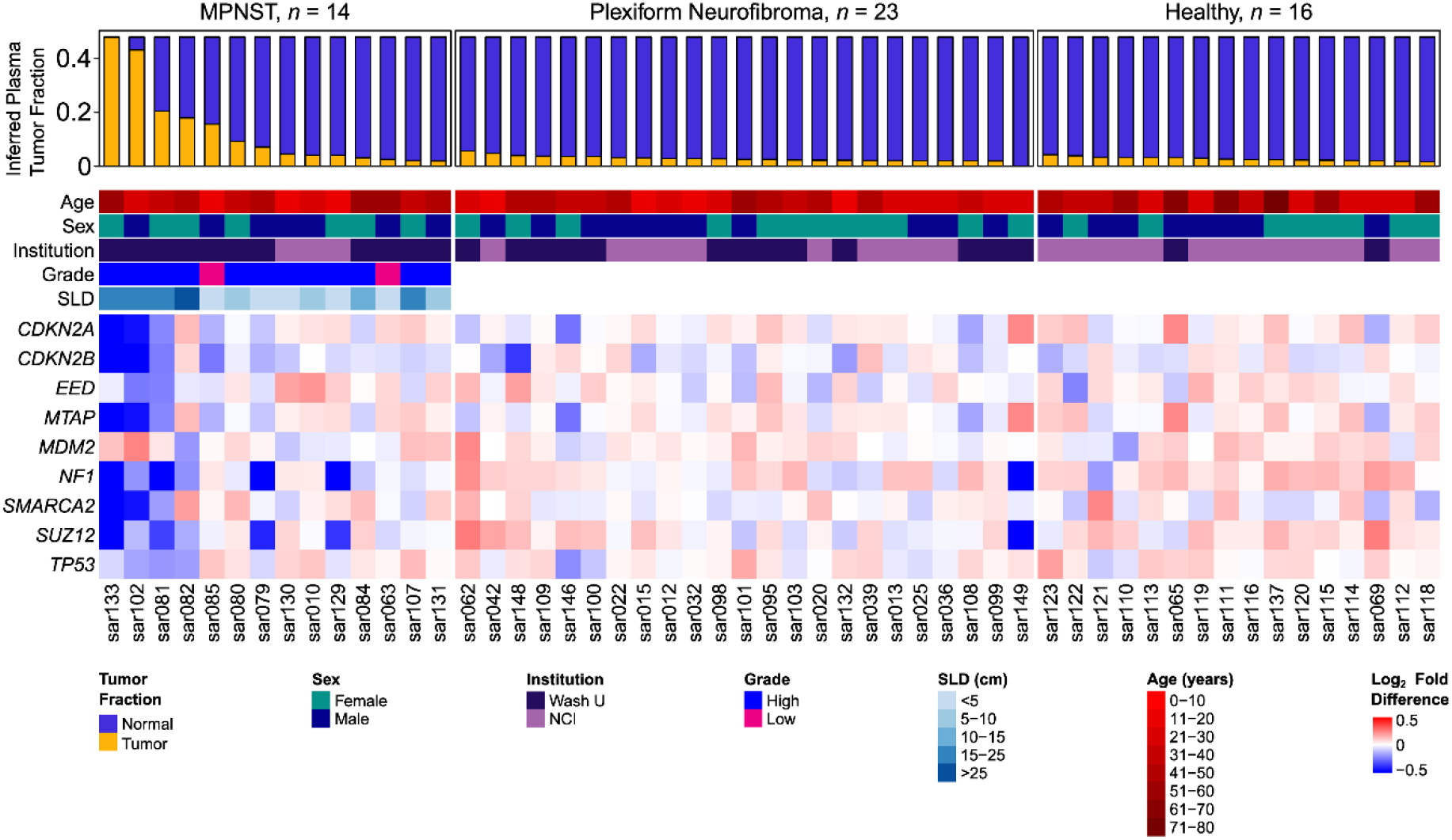
Subject characteristics and copy number alterations. Heatmap includes all 53 subjects in this study, categorized by diagnosis. Each column represents one study subject with ID labels shown below. Highest tumor fraction in plasma and important tumor and subject characteristics are displayed in the top panel. The lower panel shows copy number alterations in genes relevant to NF1 and MPNST pathogenesis, depicted as log_2_ of copy number ratio. MPNST, malignant peripheral nerve sheath tumor; NF1, neurofibromatosis type 1; SLD, sum of longest tumor diameters as determined by RECIST 1.1 criteria.

**Figure 3.**
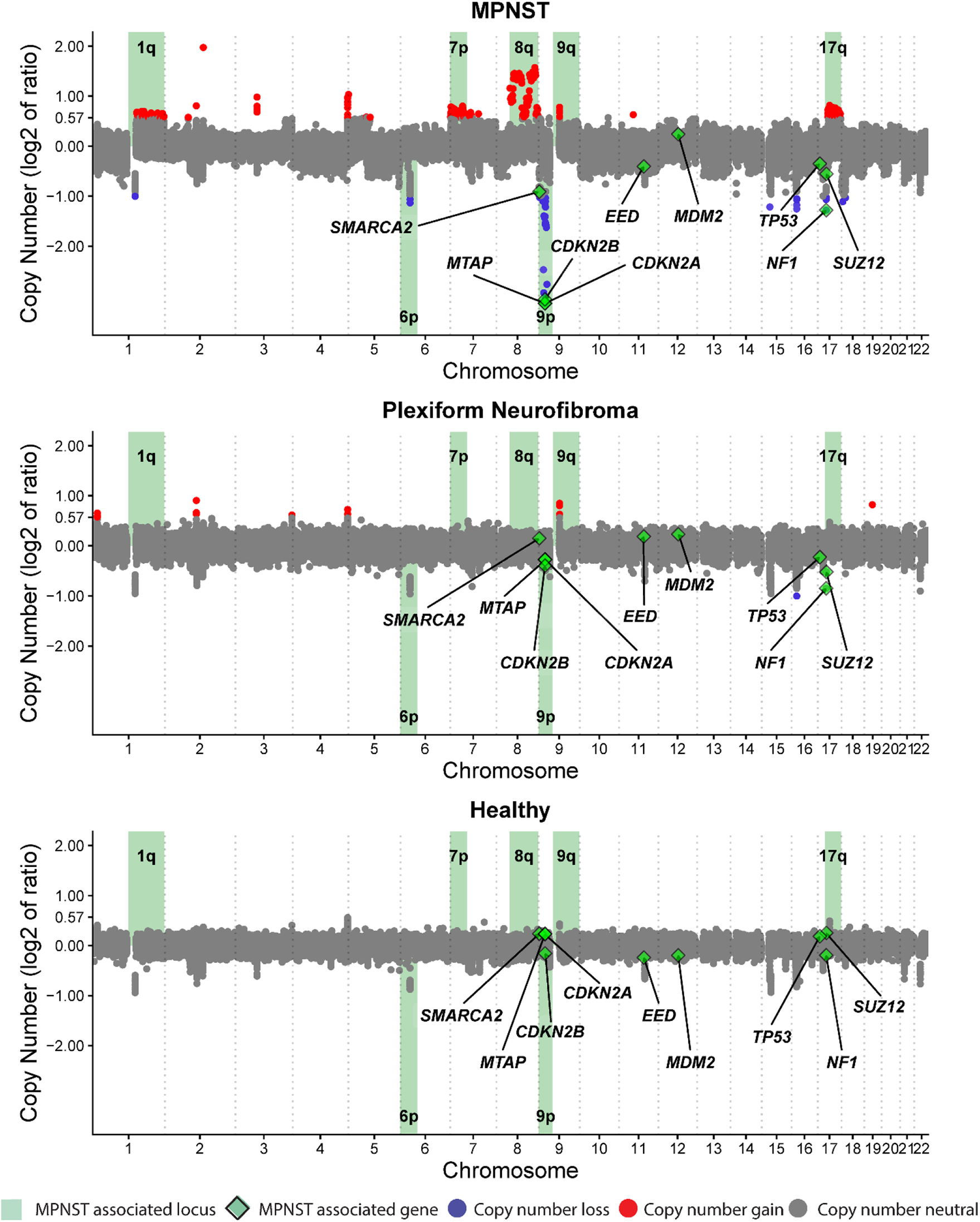
Aggregate copy number alterations measured across the genome. Plots represent plasma cell-free DNA data compiled from all blood plasma specimens from (A) MPNST (*n* = 46), (B) plexiform neurofibroma (*n* = 23) or (C) healthy donors (*n* = 16) in this study. Copy number ratios across the genome are shown on a log_2_ scale with significant gains in red, significant losses in blue, and regions without significant gain or loss depicted in gray (**Methods**). Loci highlighted in green have been previously associated with MPNST or NF1, with associated genes also labeled and depicted by green diamonds. MPNST, malignant peripheral nerve sheath tumor; PN, plexiform neurofibroma.

Given the observed copy number changes in patient plasma, we next compared genome-wide CNAs and associated tumor fractions across specimen types. For MPNST cases where tumor, leukocyte, and plasma were all available, the observed copy number aberrations were most prominent in the tumor samples, but also detected in plasma cfDNA prior to treatment, with a pattern reflective of the original tumor (**Supplementary Figure 1a**). The magnitude of these CNAs decreased in post-treatment cfDNA compared to pretreatment cfDNA, and germline samples harbored the least detectable CNAs. This trend also held for estimated tumor fractions, representing a sample’s aggregate genome-wide copy number changes. As expected, there was no such increase in tumor fraction observed in PN lesions or in cfDNA derived from PN or healthy adults.

### Plasma tumor fraction distinguishes MPNST from plexiform neurofibroma

Given that tumor-derived CNAs were detected in plasma cfDNA from MPNST patients, we next investigated the ability of plasma tumor fraction, inferred from the genome-wide copy number data, to noninvasively differentiate MPNST from PN. Plasma tumor fraction was compared between healthy controls, PN and all pretreatment MPNST samples. Strikingly, baseline cfDNA tumor fraction differentiated MPNST from both healthy (*P* = 0.0026) and PN (*P* = 0.001) subjects. PN and healthy donors did not differ significantly in tumor fraction (*P* = 1) (**Figure 4a**). Median tumor fraction levels in healthy (0.026) and PN (0.026) groups were lower than in MPNST (0.058). Comparing plasma tumor fractions between pretreatment MPNST patients and PN patients, ROC analysis further demonstrated an area under the curve of 0.83 (**Figure 4b**), which was higher with ∼0.6x ULP-WGS than ∼0.3x (**Methods; Supplementary Figure 4**). This signified the ability to accurately discriminate between MPNST and PN using only plasma tumor fraction levels derived from ULP-WGS.

Thus, utilizing a Youden’s index-optimized cut-point of 0.041, pretreatment plasma tumor fraction differentiated MPNST from PN with an area under the ROC curve of 0.83, and sensitivity of 75% and specificity of 91%, with 21 of 23 PN cases successfully classified based on pretreatment plasma tumor fraction alone (*P* = 0.001) (**Supplementary Table 5**). This compared favorably to reports of other diagnostic modalities including MRI features and image-guided core-needle biopsy (**Supplementary Table 6**). Model performance was retained in leave-one-out cross validation using a penalized regression model where overall accuracy was 75% (95% CI 66-83%), and improved to 89% with AUC of 0.89, Youden’s index-optimized sensitivity of 83% and specificity of 91% when considering the highest plasma tumor fraction measured per subject on serial timepoint analysis (**Supplementary Table 7**). In a multivariate binary logistic regression model including age, sex and institution, baseline plasma tumor fraction remained significantly associated with clinical status (*P* = 0.04) while the other covariates were not (**Supplementary Table 8**).

Validating our original fragment size selection strategy, we observed fragment size differences between clinical states as defined by the tumor fraction ROC cutpoint. Using high-tumor fraction versus low-tumor fraction groups determined by the optimal cut-point of 0.041, there was a significant difference in fragment length distributions (*D* = 0.078, *P* < 0.001 by two-sample Kolmogorov-Smirnov test) with high-plasma tumor fraction cases enriched for shorter cfDNA fragments and low-plasma tumor fraction cfDNA enriched for longer fragments (**Figure 4c**). Similarly, clinically classified MPNST patients harbored significantly shorter cfDNA fragments compared to PN patients (*D* = 0.032, *P* < 0.001) and healthy donors (**Supplementary Figure 2**). Thus, fragmentation profiles appear unique in patients with MPNST compared to those with benign PN.

**Figure 4:**
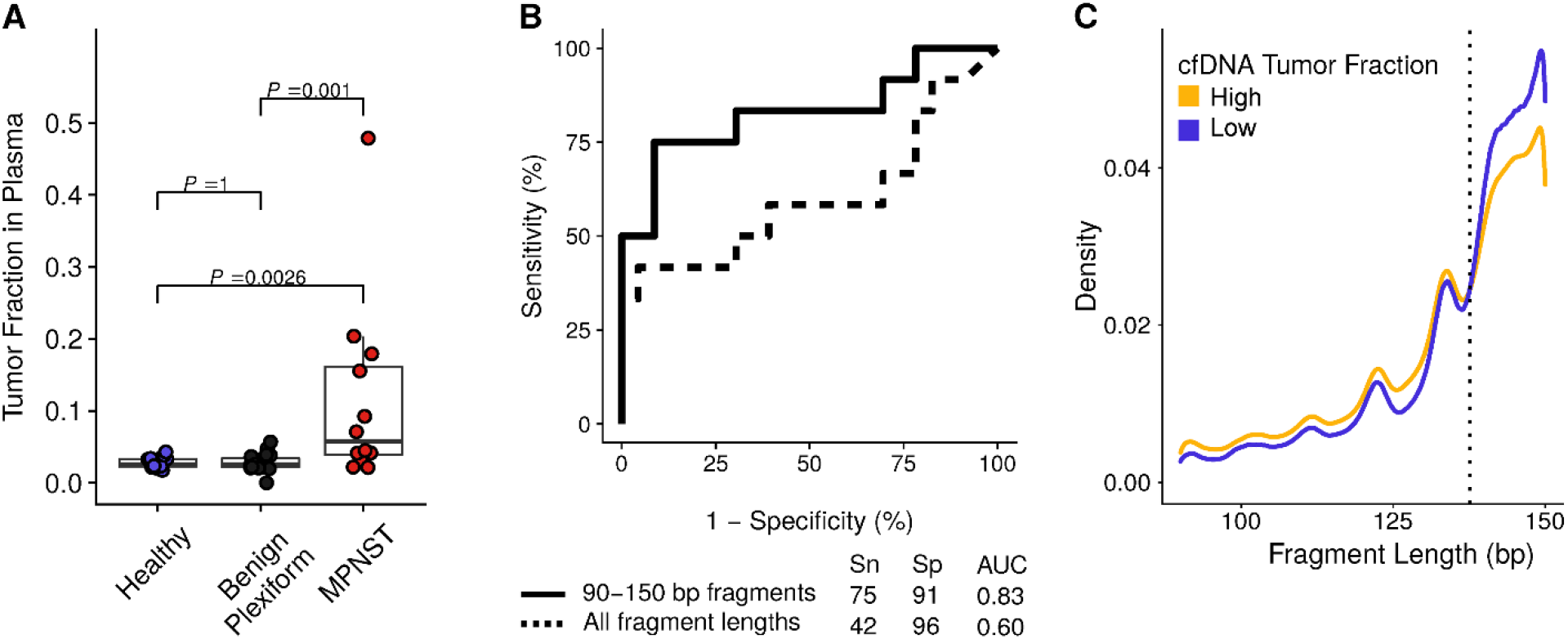
Tumor fraction in plasma stratifies subjects by diagnosis. **(A)** Tumor fraction in subjects with available pre-treatment plasma cell-free DNA (*n* = 53), stratified by clinical diagnosis, with significance assessed by the Dunn test after Kruskal-Wallis analysis of variance. **(B)** Receiver operating characteristic (ROC) curve of plasma cfDNA tumor fraction comparing pretreatment MPNST to plexiform neurofibroma patients. Solid line represents tumor fraction data derived only from 90-150 bp fragments while dotted line represents tumor fractions derived from all fragment lengths. Confusion matrix is reported separately (**Supplementary Table 5**). **(C)** Fragment length density for cell-free DNA in MPNST and PN patients (*n* = 92 samples) with high (>0.0413) versus low (<0.0413) tumor fractions in plasma as determined by the Youden’s index-optimized cutpoint from the ROC curve. The dashed line highlights an inflection in the curves with high tumor fraction samples enriched for shorter cfDNA fragment sizes (<138 bp) and low tumor fraction samples enriched for longer cfDNA fragment sizes (*D* = 0.078, *P* < 0.001 by two-sample Kolmogorov-Smirnov test). Data is shown for sequencing reads within the 90 to 150 bp in-silico size-selection range (**Methods**). AUC, area under the curve; bp, base pairs; cfDNA, cell-free DNA; MPNST, malignant peripheral nerve sheath tumor; Sn, sensitivity; Sp, specificity.

### MPNST plasma tumor fraction correlates with disease burden by imaging

Having established plasma cfDNA fragment size and tumor fraction as a specific means to classify MPNST cases noninvasively, we next investigated the relationship between plasma tumor fraction derived using our assay and radiologically measured tumor burden. Radiographic tumor burden was quantified by the sum of longest diameters (SLD) by RECIST 1.1 criteria [39] and compared to matched plasma cfDNA tumor fraction levels (**Methods**). A significantly positive correlation was observed between SLD and plasma tumor fraction (Pearson *r* = 0.387, *P* = 0.024) (**Figure 5a**). Because RECIST SLD measurements are restricted to five total lesions, two lesions per organ and do not include bony disease, SLD may underestimate tumor burden in metastatic MPNST patients [39]. Conversely, SLD may overestimate the size of malignant tissue in primary MPNST lesions which often arise from within PN, with the relative contribution of PN versus MPNST tissue to the overall lesion size difficult to accurately assess radiographically [47]. These challenges limit the ability of SLD to accurately define MPNST disease burden and may explain why the correlation of SLD to tumor fraction was not stronger.

**Figure 5:**
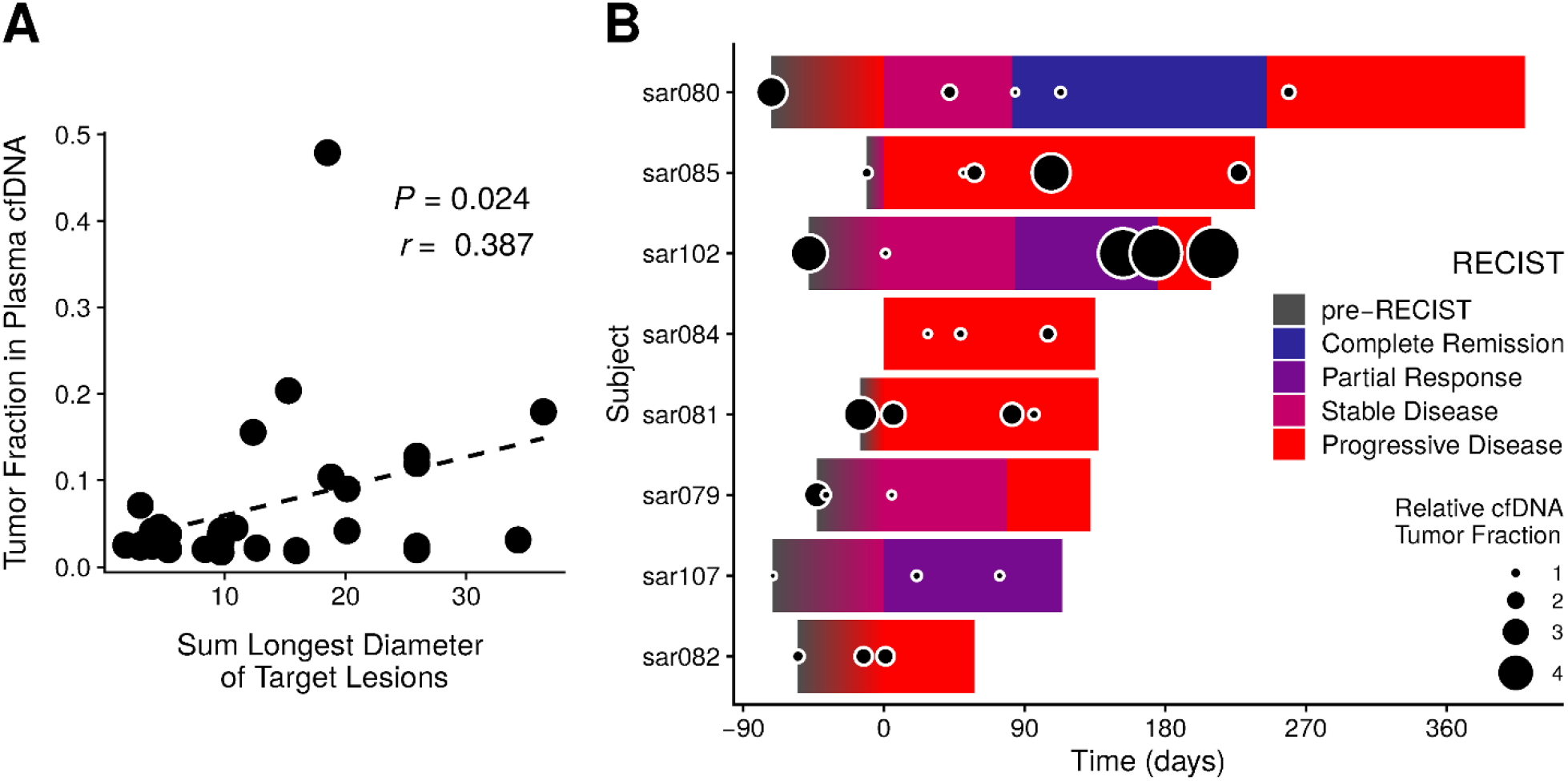
Tumor fraction in plasma correlates with imaging. **(A)** Sum of longest diameters (SLD) of target lesions for all MPNST patients are plotted against each SLD’s nearest plasma tumor fraction value (**Methods**). Pearson correlation coefficient is significant at *P* = 0.024. **(B)** Timelines of RECIST classification for MPNST patients that underwent serial monitoring (*n* = 8; **Supplementary Figure 3**). Overlaid are cfDNA tumor fraction levels, normalized per-patient to the lowest value detected on serial analysis, and then log_2_-transformed (**Methods**). cfDNA, cell-free DNA; RECIST, response evaluation criteria in solid tumors, version 1.1.

We furthermore dynamically tracked both SLD and plasma tumor fractions over time in patients with serial timepoint data (**Supplementary Table 1**). We applied RECIST 1.1 criteria [39] in these patients to classify radiographic response to therapy (**Methods**). Dynamic changes in tumor fraction typically correlated with but preceded imaging changes (**Figure 5b; Supplementary Figure 3**). We thus studied timelines of disease progression versus response for MPNST patients, comparing imaging SLD to plasma tumor fraction in the context of the specific treatments received (**Figures 6-8; Supplementary Figure 3**). For example, sar085 presented with a small recurrent lung MPNST that rapidly progressed into multiple thoracic metastases despite two lines of cytotoxic chemotherapy, but then partially responded to third-line chemotherapy (**Figure 6**). Interestingly, plasma tumor fraction levels closely tracked with SLD from CT imaging throughout this treatment course. These data suggest that plasma tumor fraction could thus be utilized to monitor treatment response.

**Figure 6.**
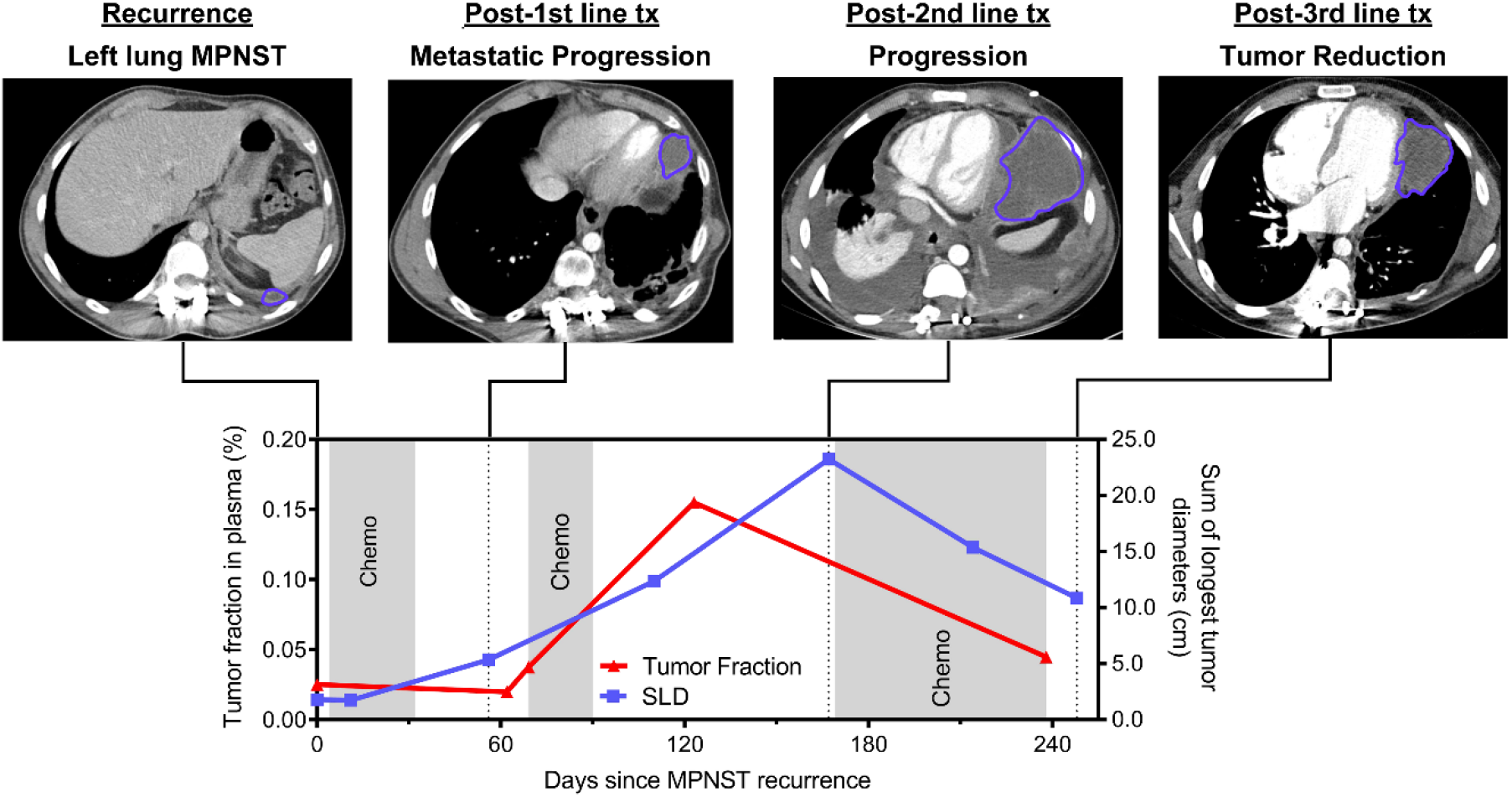
Monitoring tumor burden vignette (sar085). This patient had a high-grade thoracic MPNST recurrence that progressed rapidly through first-and second-line chemotherapy but responded to third-line chemotherapy. Tumor fraction in plasma was initially undetectable, then rapidly increased during first-and second-line chemotherapy, followed by a rapid decrease during third-line chemotherapy. This dynamic tumor fraction in plasma correlated well with the sum of longest tumor diameters (SLD) measured radiographically by RECIST 1.1 criteria. Chemo, chemotherapy; cm, centimeters; MPNST, malignant peripheral nerve sheath tumor; Tx, treatment.

There were also several instances where we observed cfDNA tumor fraction elevations anticipating and preceding corresponding SLD increases. sar80 is an illustrative example in which the patient had complete resection of a right pelvic MPNST prior to plasma cfDNA analysis (**Figure 7**). Initial plasma tumor fraction was not detected, which was consistent with our finding of no evidence of disease by imaging. Follow-up imaging again showed no evidence of disease 2.5 months later. Plasma tumor fraction rose, however, just 14 days after this most recent negative imaging study. Still, radiographic recurrence was not detected until 89 days later, suggesting that the measurement of plasma tumor fraction could be utilized as a sensitive surveillance tool for minimal residual disease detection following the completion of therapy.

**Figure 7:**
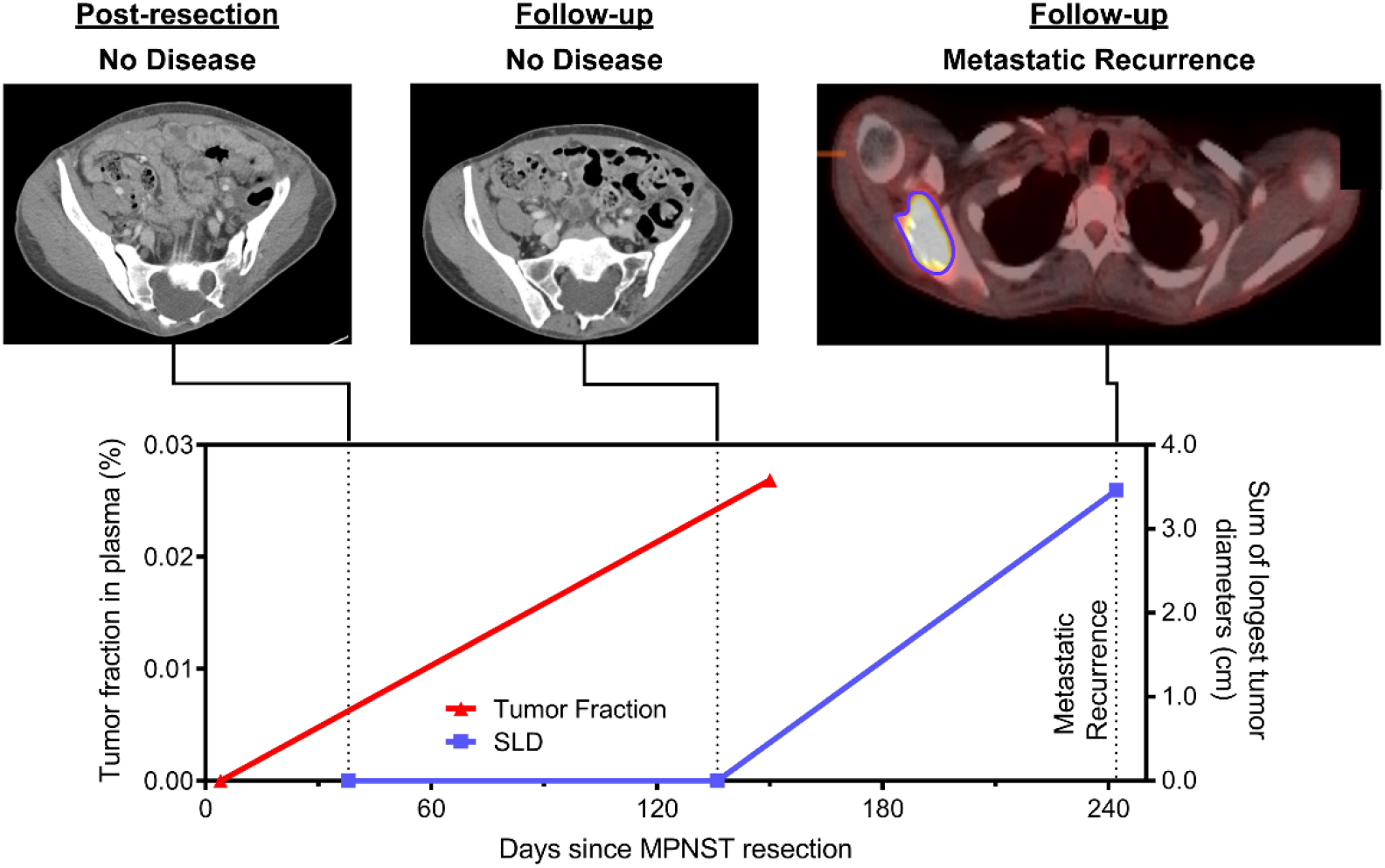
Early detection of relapse vignette (sar80). This patient previously had a high-grade pelvic MPNST that was completely resected with no evidence of residual disease after surgery. Tumor fraction in plasma was undetectable following resection but was detected 150 days later (tumor fraction = 0.021); this preceded metastatic recurrence identified on surveillance imaging by 89 days. cm, centimeters; MPNST, malignant peripheral nerve sheath tumor; SLD, sum of longest tumor diameters as determined by RECIST 1.1 criteria.

sar102 illustrates another example where cfDNA tumor fraction elevation preceded radiographic progression. This patient had metastatic MPNST with a presumed right foot primary (**Figure 8**). During and following over four months of cytotoxic chemotherapy, imaging showed a partial response with persistently decreasing SLD. Given radiographic evidence of disease control, the medical team then decided to hold cytotoxic chemotherapy for an elective lower-limb amputation with the goal of improving quality of life (pain reduction and ability to use a prosthetic limb). In contrast to SLD however, plasma tumor fraction increased 14-fold during this same interval (**Supplementary Table 1**). Following cessation of cytotoxic agents and amputation, the patient was found to have significant widespread tumor progression with the development of multiple distant metastases including brain metastases. Here, plasma tumor fraction was more consistent than serial imaging with this patient’s ultimate clinical status, and could have influenced the decision to hold chemotherapy in order to perform an elective, palliative amputation. This illustrates the potential utility of plasma cfDNA tumor fraction analysis for dynamically monitoring disease, anticipating progression, and influencing clinical decision-making.

**Figure 8:**
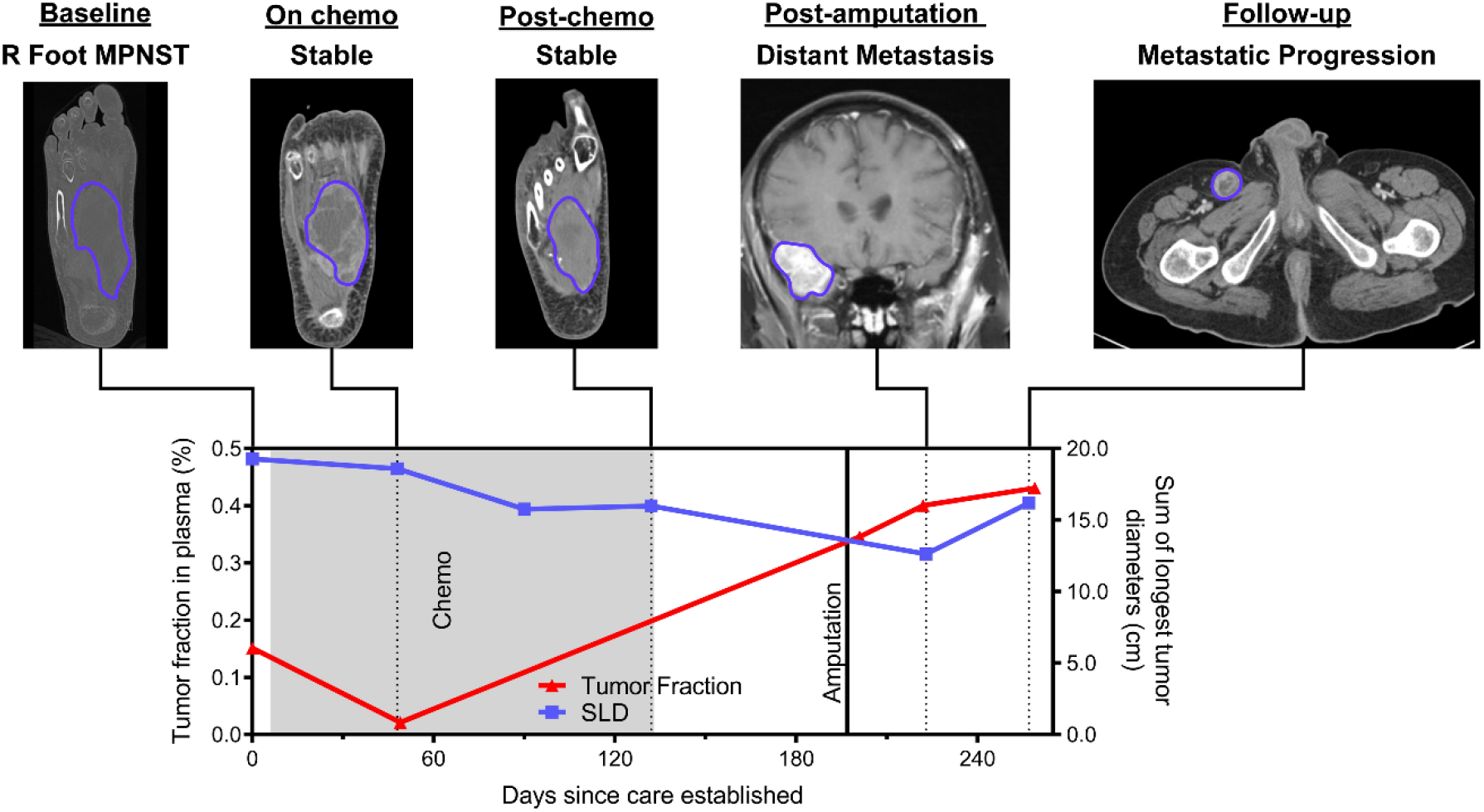
Clinical decision-making vignette (sar102). This patient had a high-grade MPNST in the right foot that was stable by imaging and therefore had chemotherapeutic agents held in order to undergo an elective lower limb amputation for improved quality of life. Tumor fraction in plasma, however, increased during this pre-surgical time period, consistent with progressive metastatic disease which became apparent on imaging shortly after surgery. The patient would not have had chemotherapy held to undergo lower limb amputation had there been earlier evidence of progressive metastatic disease. Chemo, chemotherapy; cm, centimeters; MPNST, malignant peripheral nerve sheath tumor; SLD, sum of longest tumor diameters as determined by RECIST 1.1 criteria.

## Discussion

In this multi-institutional cohort study, we performed fragment size analysis and ultra-low-pass whole genome sequencing (ULP-WGS) of cfDNA to noninvasively detect genome-wide CNAs and derive tumor fraction in plasma, which we used to differentiate between MPNST and PN patients. To our knowledge, this is the first demonstration of liquid biopsy to distinguish between malignant and pre-malignant solid tumor in the setting of a cancer predisposition syndrome. Specifically, we observed that patients with MPNST harbor a unique cfDNA fragmentation profile and have significantly greater tumor genomic instability evident in plasma compared to PN patients. We also demonstrated that cfDNA analysis can be used to dynamically track treatment response in MPNST patients, potentially with greater precision than standard cross-sectional imaging.

MPNST are aggressive soft tissue sarcomas that can be difficult to distinguish from their benign precursors, illustrating the need for new testing modalities for better disease detection and surveillance. Our data illustrate several important points. First, we accurately identified copy number-altered genomic loci characteristic of malignant transformation from PN using ultra-low-pass WGS of cfDNA (**Figures 2 and 3**) [32,43]. Specifically, loss of *CDKN2A/B, SMARCA2*, and *SUZ12* were commonly found only in MPNST plasma samples, while loss of *NF1* was observed in both MPNST and PN plasma, but not healthy controls. When paired tumor was available, plasma cfDNA-detected CNAs recapitulated tissue patterns of genomic instability (**Supplementary Figure 1**). Together, these data suggest that even at low sequencing coverage, the genomic features of both NF1 and of progression from PN to MPNST are detectable in affected patients’ plasma.

Here we show that plasma tumor fraction derived from genome-wide CNAs, without applying prior knowledge of patient-specific mutational profiles, differentiated MPNST from PN with high specificity (91%) and moderate sensitivity (75%) pre-treatment (**Figure 4b**) and high specificity (91%) and sensitivity (83%) when measured serially (**Supplementary Table 7**), suggesting that cfDNA tumor fraction could be a valuable adjunct to aid in monitoring patients with PN with the goal of early cancer detection. Currently, malignant transformation in NF1 patients is difficult to screen for due to overlapping clinical symptoms and radiographic findings that are also associated with benign PN [48,49]. Reflective of this, current standard practice for PN surveillance is to obtain imaging only when clinically indicated. Moreover, clinical surveillance for symptoms such as lesion-associated pain have a low specificity for identifying MPNST on subsequent workup [48,50,51]. Furthermore, the lack of reliable radiographic characteristics using standard sequences that can be replicated across institutions has contributed to overall limited sensitivity of MRI (62.5-84%) and specificity of FDG-PET (52.2-83%) for MPNST detection [20,21,52].

Still, it will be important to fully consider state-of-the-art imaging results, such as anatomic MRI features [53] and integrate them with plasma cfDNA results in the future to potentially maximize combined modality performance, which we suggest may be possible by coupling the high sensitivity but relatively low specificity of advanced MRI features with our highly specific liquid biopsy assay (**Supplementary Table 6**). Indeed, in current practice, confirmation of MPNST identified by clinical/imaging suspicion is usually attempted by solid tumor biopsy, which can be exquisitely painful given the peripheral nerve site, is often technically difficult due to lesions’ propensity for localizing to viscera and the retroperitoneum, and is associated with serious complications including nerve palsy and dissemination of malignant tumor cells [53]. Additionally, biopsy is not without diagnostic caveat, as the development of MPNST from within PN lesions causes sampling bias with image-guided biopsies shown to result in low negative predictive value (NPV), with 50% of NF1 patients diagnosed with PN on image-guided core-needle biopsy being subsequently reclassified as MPNST following surgical resection in one retrospective study [19]. Unlike image-guided tumor biopsy, our liquid biopsy approach to measure tumor fraction reflects chromosomal instability throughout the body, limiting the potential for sampling bias.

Previous studies have demonstrated the ability of liquid biopsy testing to distinguish patients with cancer from healthy individuals [26,54-57]. Here we significantly extend this body of work to show that a liquid biopsy test can detect malignant transformation in the context of hereditary cancer predisposition, distinguishing patients with a malignant solid tumor from those with its benign precursor lesion. We also build upon prior literature showing that fragmentation profiles and lengths of cell-free DNA from cancer patients are distinct from those in healthy donors [26,28-30], showing for the first time that malignancy-associated cfDNA is significantly shorter than its benign-associated counterpart (**Figure 4c, Supplementary Figure 2**). We additionally demonstrate that increasing sequencing coverage to ∼0.6x improves the sensitivity of our ULP-WGS-based assay (**Supplementary Figure 4**).

Our study’s moderate pretreatment sensitivity of our cfDNA technology for detecting MPNST versus PN, which we showed was 75% with 91% specificity compares favorably to landmark early cancer detection studies such as the ones from Grail [56], Thrive [55], and Stanford [54], which reported sensitivities of ∼30-50% with ∼95-99% specificity for common stage 1-2 solid tumor malignancies. Indeed, when we set the specificity of our assay to 100%, we still achieved a comparable sensitivity of 50% pretreatment and 58% when considering the highest tumor fraction on serial timepoint analysis (**Supplementary Table 7**). Overall, our plasma cfDNA assay was able to robustly distinguish MPNST from its PN precursor, and should be tested in the setting of other hereditary cancer predisposition syndromes in the future. We nonetheless plan to enhance the technology we present here in order to boost sensitivity, for example through deep learning following future generation of much larger cfDNA datasets, and by pairing our fragmentation-and WGS-based method with targeted deep sequencing and methylation analysis.

We also show that cfDNA tumor fraction dynamics appear to anticipate and track with disease burden in MPNST patients. CNAs in plasma cfDNA have been previously shown to correlate with radiographic burden of disease in established cancer patients [58]. Our study, similarly, showed significant correlation between plasma tumor fraction and radiographic tumor burden (**Figure 5a**). Additionally, for individual patients with serial plasma samples and serial imaging studies, dynamic changes in cfDNA tumor fraction predicted changes in tumor burden and disease state (**Figure 5b-8; Supplementary Figure 3**). These findings highlight the potential for ULP-WGS surveillance in NF1, not only to distinguish between pre-malignant and malignant tumors, but also to serve as a real-time biomarker to track treatment response and to improve detection of minimal residual disease (MRD) following local disease control. Multi-institutional prospective validation and evaluation in cfDNA-guided interventional trials is warranted.

Limitations of our study include a modest MPNST cohort size. Indeed, landmark publications on MPNST genomics comprise of whole genome sequencing (WGS)/whole exome sequencing (WES) cohorts ranging from 7 to 15 patients [43,45], comparable to our MPNST cohort size obtained from prospectively enrolling from two major NF1 referral centers. Still, due to our modest study size, we were unable to power a held-out validation cohort (see **Methods**). To address this, we validated our data using a leave-one-out cross-validation framework which we show retains an overall accuracy of 75%. Ultimately, a much larger multi-institutional collaboration will be required to validate our method and the MPNST vs. PN tumor fraction cutpoint. A second important limitation of this study was inconsistent testing and treatment protocols across cohort subjects. Imaging data was at clinician discretion using a variety of modalities and timepoints. Germline NF1 testing was not conducted in all patients, and not all patients met NIH NF1 criteria. While lack of uniform clinical care likely reduced our ability to differentiate disease states, it also reflects real-world diversity in treatment expected in any large MPNST cohort outside of a dedicated prospective trial.

In conclusion, our findings suggest that cfDNA fragment analysis followed by ULP-WGS can noninvasively detect MPNST, and distinguish it from its benign precursor lesion in NF1 patients. To our knowledge, these results here represent the first evidence of a liquid biopsy test to capably differentiate between malignant and pre-malignant tumors in a heritable cancer predisposition syndrome. Application of this liquid biopsy technology has the potential to adjudicate equivocal imaging, serve as a MRD and treatment response biomarker, and most importantly facilitate the early detection of MPNST. These advances are critical for improving the substantial morbidity and mortality associated with these aggressive tumors in patients with this common cancer predisposition syndrome.

## Data Availability

Data will be publicly available following print publication of the study.

https://doi.org/10.7303/syn23651229

## Acknowledgements

We are grateful to the patients and families involved in this study, to the clinical research team for collection of samples and clinical data, to the Washington University Neurofibromatosis (NF) Center, and to the NCI Center for Cancer Research Intramural Research Program. We also thank D. Gutmann, T. Ley and A. Newman for providing critical feedback on the manuscript. This work was supported by grants from the Children’s Cancer Foundation NextGen Award (R.T.S.), the National Institute of General Medical Sciences (5T32GM007067 supporting P.A.J.), the NCI Center for Cancer Research Intramural Research Program (1ZIABC011722-04 supporting R.T.S, J.F.S., and 1ZIABC010801-13 supporting B.C.W.), the Francis S. Collins Scholars Program in Neurofibromatosis Clinical and Translational Research (A.C.H.), the St. Louis Men’s Group Against Cancer (A.C.H.), the Washington University Alvin J. Siteman Cancer Research Fund (A.A.C.), the National Cancer Institute (1K08CA238711-01 to A.A.C.), the Cancer Research Young Investigator Award (A.A.C.), and the V Foundation for Cancer Research (A.A.C.). The funders had no role in study design, data collection and analysis, decision to publish, or preparation of the manuscript. This study utilized the computational resources of the McDonnell Genome Institute at Washington University, and the High Performance Computing Biowulf cluster at the National Institutes of Health. Images from BioRender were used to create Figure 1.

## Supplementary Figures and Tables

**Supplementary Figure 1.**
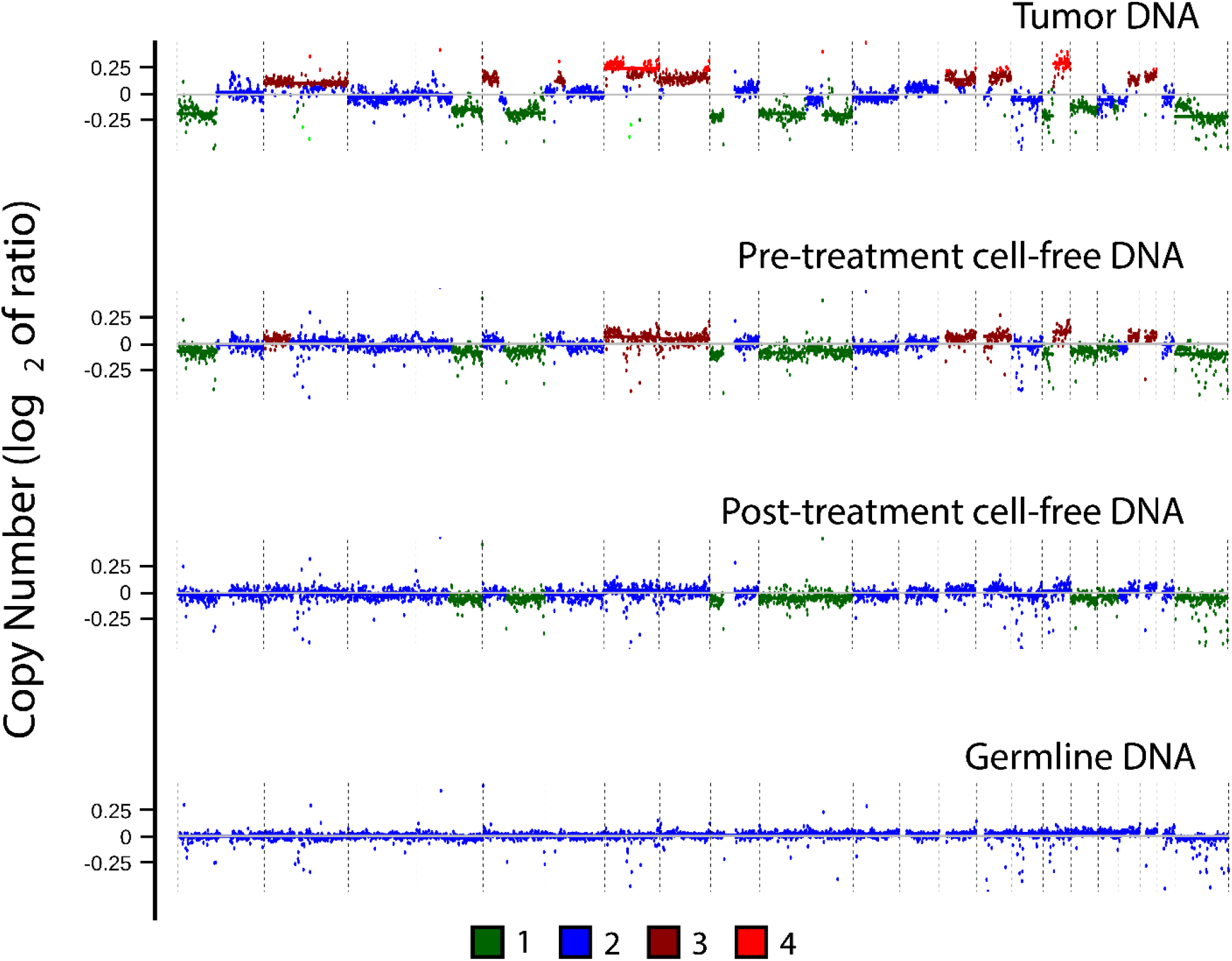
Genome-wide copy number alterations by specimen type. Genome-wide copy number alterations assessed in four specimen types from a single MPNST patient (sar081): tumor tissue DNA, pretreatment blood plasma cfDNA, post-treatment cfDNA, and germline DNA from pretreatment peripheral blood mononuclear cells. Log_2_ of copy number ratio is shown across the genome. Color scale depicts estimated copy number within the tumor fraction as determined by ichorCNA (**Methods**).

**Supplementary Figure 2.**
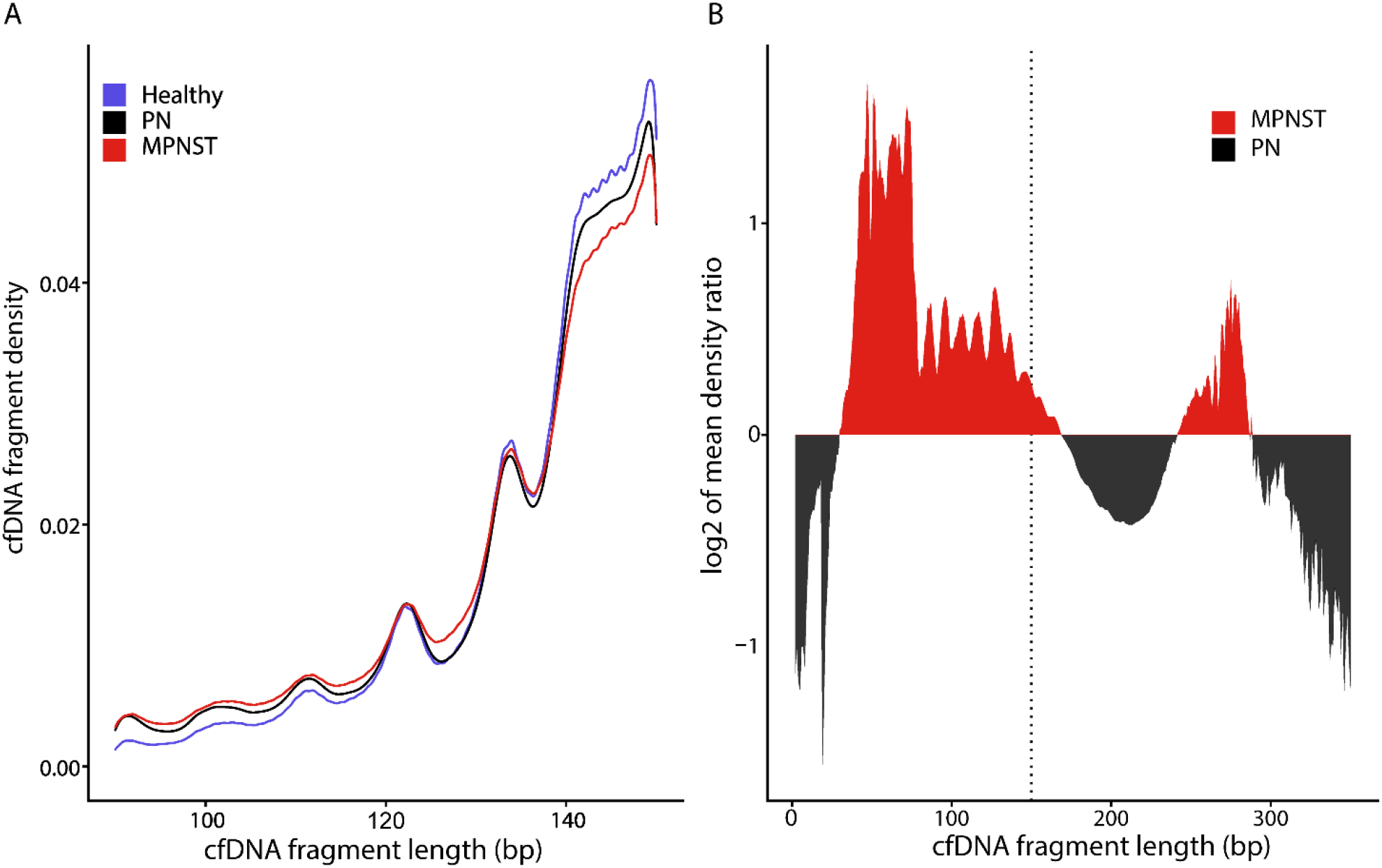
Plasma cell-free DNA fragment sizes in MPNST patients are shorter than in plexiform neurofibroma patients or healthy controls. (**A**) Fragment size distributions of cfDNA from healthy donors, PN and MPNST patients (**Methods**). cfDNA fragment sizes in MPNST patients were significantly shorter than from PN patients (*D* = 0.032, *P* < 0.001) or healthy donors (*D* = 0.062, *P* < 0.001) by two-sample Kilmogorov-Smirnov testing. (**B**) Log_2_ ratio of the differences in cfDNA fragment sizes from patients with MPNST versus PN with the dashed line indicating the upper boundary used for in-silico size selection (150 bp). For panel A, all plasma samples in the study were analyzed (16 healthy, 23 PN, 46 MPNST), and in panel B, all PN (*n* = 23) and MPNST (*n* = 46) plasma samples were analyzed. bp, base pairs; cfDNA, cell-free DNA; MPNST, malignant peripheral nerve sheath tumor; PN, plexiform neurofibroma

**Supplementary Figure 3.**
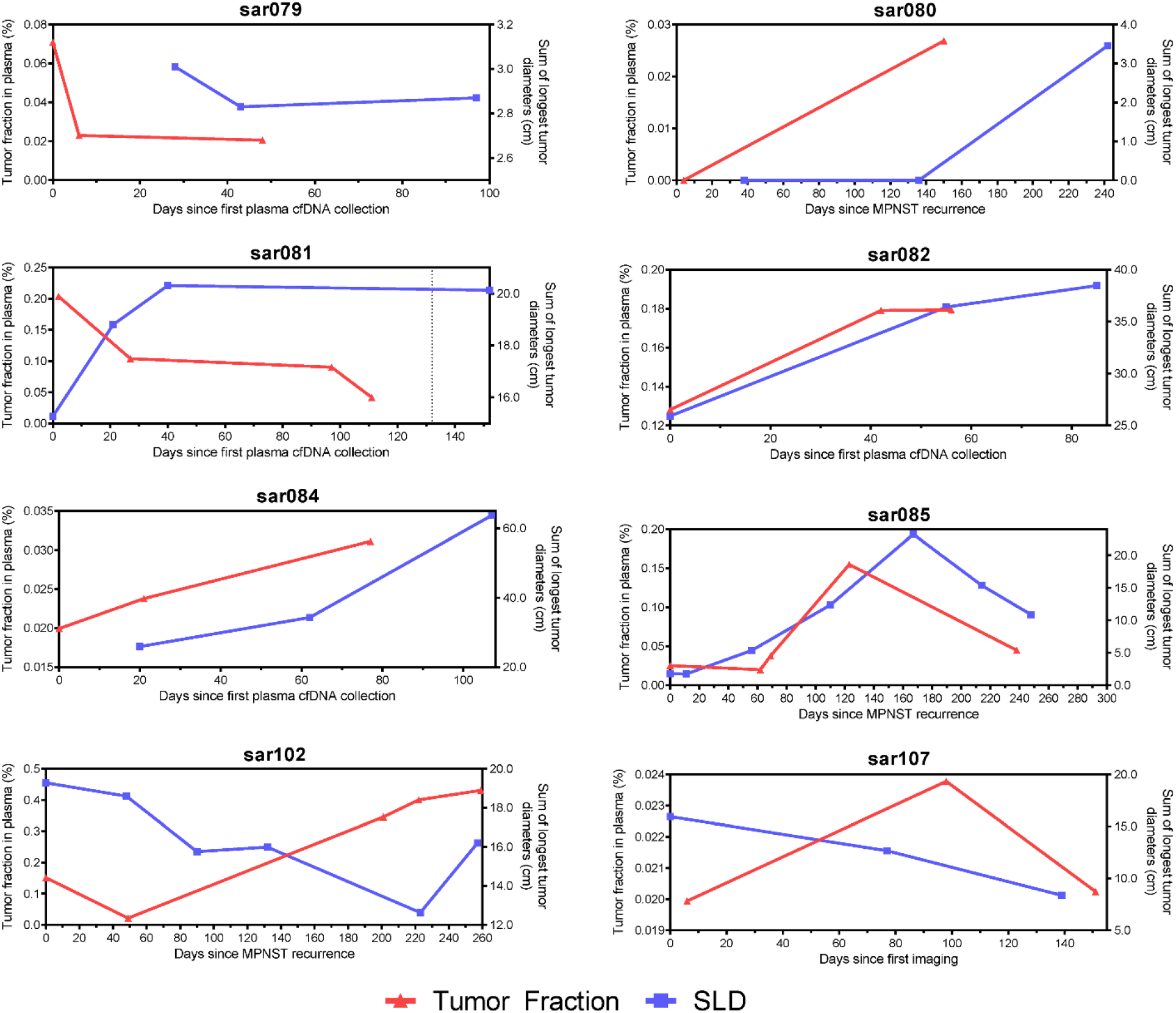
Plasma tumor fraction changes dynamically with imaging in MPNST patients. Overlaid plots of cfDNA tumor fraction (red) and SLD (blue) for MPNST patients tracked with serial plasma analysis (see also **Figure 5b**). cfDNA, cell-free DNA; SLD, sum of longest tumor diameters as determined by RECIST 1.1 criteria.

**Supplementary Figure 4.**
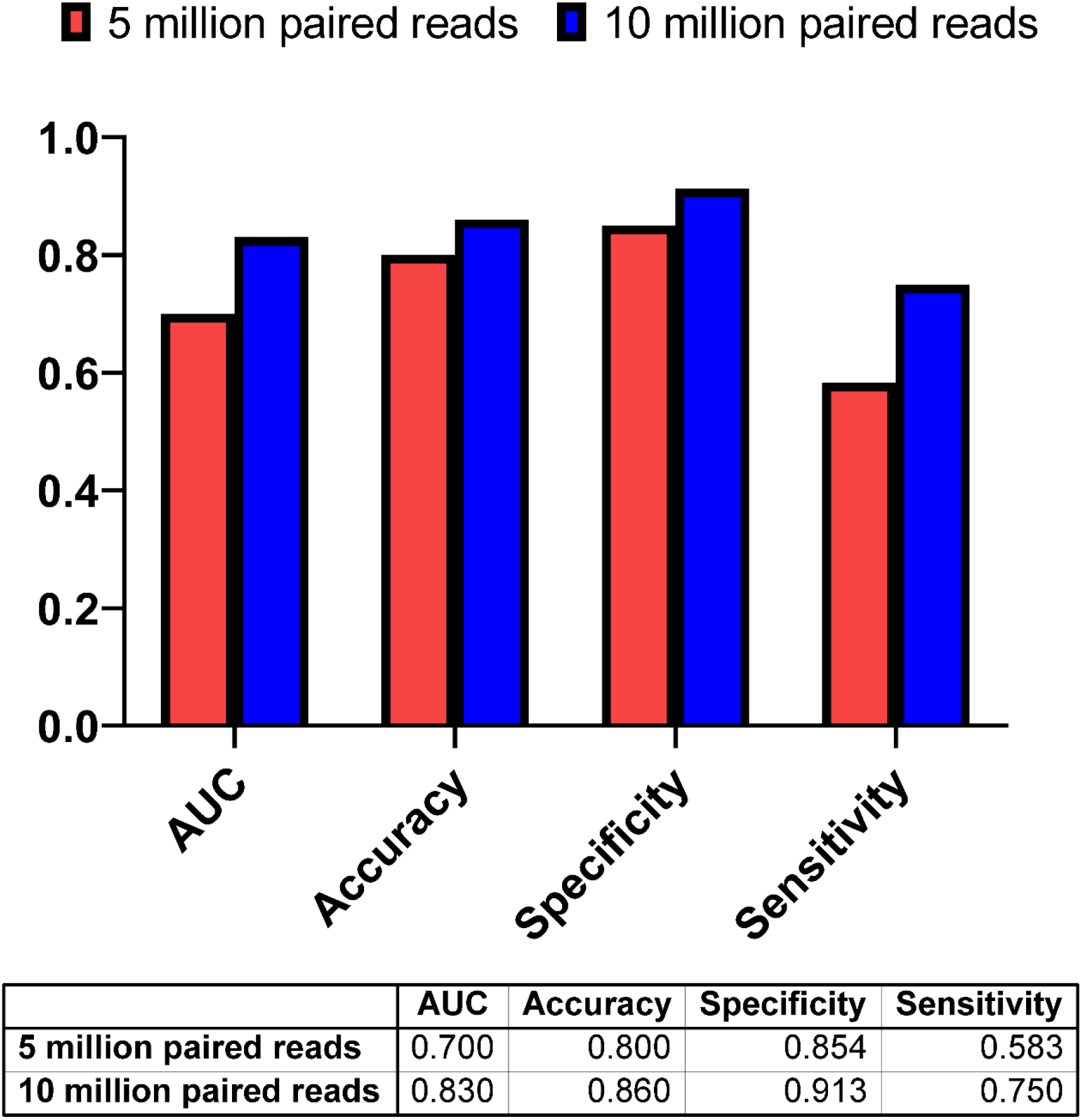
Higher plasma ULP-WGS sequencing depth improves the ability to differentiate MPNST from PN. Comparison of receiver operating characteristics (ROC) summary statistics from 5 million paired reads (∼0.3x coverage) versus 10 million paired reads (∼0.6x coverage) followed by size selection of 90-150 bp fragments. AUC, area under the curve; bp, base pairs; ROC, receiver operating characteristic.

**Supplementary Table 1. Details of all sequencing libraries in this study**.

**Supplementary Table 2. MPNST patient characteristics**.

**Supplementary Table 3. Plexiform neurofibroma patient characteristics**.

**Supplementary Table 4. Healthy adult donor characteristics**.

**Supplementary Table 5. Confusion matrix comparing cfDNA assay performance to pre-treatment MPNST vs. PN diagnosis**.

**Supplementary Table 6. Comparison of diagnostic tests’ ability to distinguish MPNST from PN**.

**Supplementary Table 7. Assay performance with alternative ROC cutpoints and timepoints**.

**Supplementary Table 8. Multivariate logistic regression predicting MPNST vs. plexiform neurofibroma clinical status**.

### Abbreviations

AN: atypical neurofibroma
CNA: copy number alteration
cfDNA: cell-free DNA
ctDNA: circulating tumor DNA
FFPE: formalin-fixed paraffin-embedding
IQR: interquartile range
NCI: National Cancer Institute
NF1: neurofibromatosis type 1
PBMC: peripheral blood mononuclear cell
PN: plexiform neurofibroma
MPNST: malignant peripheral nerve sheath tumor
MRD: minimal residual disease
ROC: receiver operating characteristic
SLD: sum of longest diameters
ULP-WGS: ultra-low-pass whole genome sequencing
WES: whole exome sequencing
WGS: whole genome sequencing
WUSTL: Washington University in Saint Louis

## Notes

### Competing Interest Statement

I have read the journal's policy and the authors of this manuscript have the following
competing interests: A.A.C. has patent filings related to cancer biomarkers. A.A.C. has
served as a consultant/advisor to Roche, Tempus, Geneoscopy, NuProbe, Fenix
Group International and Guidepoint, and receives research support from Roche. A.A.C.
is an inventor of intellectual property licensed to Biocognitive Labs, has stock options in
Geneoscopy, and ownership interests in Droplet Biosciences. F.Q. has stocks:
Centene. No potential conflicts of interest were disclosed by the other authors.

### Funding Statement

This work was supported by grants from the Childrens Cancer Foundation NextGen Award (R.T.S.), the National Institute of General Medical Sciences (5T32GM007067 supporting P.A.J.), the NCI Center for Cancer Research Intramural Research Program (1ZIABC011722-04 supporting R.T.S, J.F.S., and 1ZIABC010801-13 supporting B.C.W.), the Francis S. Collins Scholars Program in Neurofibromatosis Clinical and Translational Research (A.C.H.), the St. Louis Mens Group Against Cancer (A.C.H.), the Washington University Alvin J. Siteman Cancer Research Fund (A.A.C.), the National Cancer Institute (1K08CA238711-01 to A.A.C.), the Cancer Research Young Investigator Award (A.A.C.), and the V Foundation for Cancer Research (A.A.C.). The funders had no role in study design, data collection and analysis, decision to publish, or preparation of the manuscript.

### Author Declarations

NCI protocol NCT01109394, NIH Intramural IRB identifier 10C0086; NCI protocol NCT00924196, NIH Intramural IRB identifier 08C0079; WUSTL protocol NCT04354064, Washington University in St. Louis School of Medicine Human Research Protection Office IRB identifiers 201903142 and 201203042

### Summary of Updates

The manuscript has been updated to reflect a technical improvement in the method. The 2017 Nature Communications paper first describing ichorCNA (Adalsteinsson et al., 2017) reported that at least 10% tumor content can be quantified from cfDNA sequenced with 0.1x coverage. To achieve a lower limit of detection in the setting of cancer vs. pre-cancer detection, we sequenced our libraries more deeply to ~0.2-6x coverage, although we did not previously do a formal analysis correlating sequencing coverage with assay performance. We have now done this analysis, comparing ~0.3x coverage (5 million read-pairs) to ~0.6x coverage (10 million read-pairs) plasma WGS, which reveals improved assay performance with higher sequencing depth, as shown in new Supplemental Figure 4 in the updated manuscript. We were able to apply ~0.6x coverage NGS to our original full MPNST cohort (n=14 subjects, n=46 samples), healthy donors (n=16 subjects, n=16 samples), and an adequately powered subset of the PN cohort (n=23 subjects, n=23 samples) per our pre-study sample size calculation (see Methods). The ability of deeper sequencing to optimize tumor fraction inference following low-pass WGS of plasma cfDNA for cancer vs. pre-cancer detection is a new technical innovation which we now highlight in our updated Discussion section.

## References

1. Evans DG, Baser ME, McGaughran J, Sharif S, Howard E, Moran A. Malignant peripheral nerve sheath tumours in neurofibromatosis 1. J Med Genet. 2002;39(5):311-4. Epub 2002/05/16. doi: 10.1136/jmg.39.5.311. PubMed PMID: 12011145; PubMed Central PMCID: PMCPMC1735122.

2. Peltonen J, Jaakkola S, Lebwohl M, Renvall S, Risteli L, Virtanen I, et al. Cellular differentiation and expression of matrix genes in type 1 neurofibromatosis. Lab Invest. 1988;59(6):760-71. Epub 1988/12/01. PubMed PMID: 2462129.

3. Serra E, Rosenbaum T, Winner U, Aledo R, Ars E, Estivill X, et al. Schwann cells harbor the somatic NF1 mutation in neurofibromas: evidence of two different Schwann cell subpopulations. Hum Mol Genet. 2000;9(20):3055-64. Epub 2000/12/15. doi: 10.1093/hmg/9.20.3055. PubMed PMID: 11115850.

4. Basu TN, Gutmann DH, Fletcher JA, Glover TW, Collins FS, Downward J. Aberrant regulation of ras proteins in malignant tumour cells from type 1 neurofibromatosis patients. Nature. 1992;356(6371):713-5. Epub 1992/04/23. doi: 10.1038/356713a0. PubMed PMID: 1570015.

5. Kahen EJ, Brohl A, Yu D, Welch D, Cubitt CL, Lee JK, et al. Neurofibromin level directs RAS pathway signaling and mediates sensitivity to targeted agents in malignant peripheral nerve sheath tumors. Oncotarget. 2018;9(32):22571-85. Epub 2018/06/02. doi: 10.18632/oncotarget.25181. PubMed PMID: 29854299; PubMed Central PMCID: PMCPMC5978249.

6. Nur Ekms, Varga M, Maruta H. The GTPase-activating NF1 fragment of 91 amino acids reverses v-Ha-Ras-induced malignant phenotype. J Biol Chem. 1993;268(30):22331-7. Epub 1993/10/25. PubMed PMID: 8226742.

7. Miettinen MM, Antonescu CR, Fletcher CDM, Kim A, Lazar AJ, Quezado MM, et al. Histopathologic evaluation of atypical neurofibromatous tumors and their transformation into malignant peripheral nerve sheath tumor in patients with neurofibromatosis 1-a consensus overview. Hum Pathol. 2017;67:1-10. Epub 2017/05/30. doi: 10.1016/j.humpath.2017.05.010. PubMed PMID: 28551330; PubMed Central PMCID: PMCPMC5628119.

8. Maertens O, Brems H, Vandesompele J, De Raedt T, Heyns I, Rosenbaum T, et al. Comprehensive NF1 screening on cultured Schwann cells from neurofibromas. Hum Mutat. 2006;27(10):1030-40. Epub 2006/08/31. doi: 10.1002/humu.20389. PubMed PMID: 16941471.

9. Kim A, Stewart DR, Reilly KM, Viskochil D, Miettinen MM, Widemann BC. Malignant Peripheral Nerve Sheath Tumors State of the Science: Leveraging Clinical and Biological Insights into Effective Therapies. Sarcoma. 2017;2017:7429697. Epub 2017/06/09. doi: 10.1155/2017/7429697. PubMed PMID: 28592921; PubMed Central PMCID: PMCPMC5448069.

10. Uusitalo E, Rantanen M, Kallionpaa RA, Poyhonen M, Leppavirta J, Yla-Outinen H, et al. Distinctive Cancer Associations in Patients With Neurofibromatosis Type 1. J Clin Oncol. 2016;34(17):1978-86. Epub 2016/03/02. doi: 10.1200/JCO.2015.65.3576. PubMed PMID: 26926675.

11. Ducatman BS, Scheithauer BW, Piepgras DG, Reiman HM, Ilstrup DM. Malignant peripheral nerve sheath tumors. A clinicopathologic study of 120 cases. Cancer. 1986;57(10):2006-21. Epub 1986/05/15. doi: 10.1002/1097-0142(19860515)57:10<2006::aid-cncr2820571022>3.0.co;2-6. PubMed PMID: 3082508.

12. Miller DT, Freedenberg D, Schorry E, Ullrich NJ, Viskochil D, Korf BR, et al. Health Supervision for Children With Neurofibromatosis Type 1. Pediatrics. 2019;143(5). Epub 2019/04/24. doi: 10.1542/peds.2019-0660. PubMed PMID: 31010905.

13. Zhang X, Murray B, Mo G, Shern JF. The Role of Polycomb Repressive Complex in Malignant Peripheral Nerve Sheath Tumor. Genes (Basel). 2020;11(3). Epub 2020/03/19. doi: 10.3390/genes11030287. PubMed PMID: 32182803; PubMed Central PMCID: PMCPMC7140867.

14. Prudner BC, Ball T, Rathore R, Hirbe AC. Diagnosis and management of malignant peripheral nerve sheath tumors: Current practice and future perspectives. Neurooncol Adv. 2020;2(Suppl 1):i40-i9. Epub 2020/07/10. doi: 10.1093/noajnl/vdz047. PubMed PMID: 32642731; PubMed Central PMCID: PMCPMC7317062.

15. Gachiani J, Kim D, Nelson A, Kline D. Surgical management of malignant peripheral nerve sheath tumors. Neurosurg Focus. 2007;22(6):E13. Epub 2007/07/07. PubMed PMID: 17613204.

16. Compston A. The Neurofibromatoses. A pathogenetic and clinical overview. J Neurol Neurosurg Psychiatry. 1994;57(10):1301-. PubMed PMID: PMC485524.

17. Friedman JM, Birch PH. Type 1 neurofibromatosis: a descriptive analysis of the disorder in 1,728 patients. Am J Med Genet. 1997;70(2):138-43. Epub 1997/05/16. doi: 10.1002/(sici)1096-8628(19970516)70:2<138::aid-ajmg7>3.0.co;2-u. PubMed PMID: 9128932.

18. Waggoner DJ, Towbin J, Gottesman G, Gutmann DH. Clinic-based study of plexiform neurofibromas in neurofibromatosis 1. Am J Med Genet. 2000;92(2):132-5. Epub 2000/05/08. PubMed PMID: 10797438.

19. Graham DS, Russell TA, Eckardt MA, Motamedi K, Seeger LL, Singh AS, et al. Oncologic Accuracy of Image-guided Percutaneous Core-Needle Biopsy of Peripheral Nerve Sheath Tumors at a High-volume Sarcoma Center. Am J Clin Oncol. 2019;42(10):739-43. Epub 2019/08/23. doi: 10.1097/COC.0000000000000591. PubMed PMID: 31436746.

20. Ahlawat S, Blakeley JO, Rodriguez FJ, Fayad LM. Imaging biomarkers for malignant peripheral nerve sheath tumors in neurofibromatosis type 1. Neurology. 2019;93(11):e1076-e84. Epub 2019/08/10. doi: 10.1212/WNL.0000000000008092. PubMed PMID: 31395668.

21. Benz MR, Czernin J, Dry SM, Tap WD, Allen-Auerbach MS, Elashoff D, et al. Quantitative F18-fluorodeoxyglucose positron emission tomography accurately characterizes peripheral nerve sheath tumors as malignant or benign. Cancer. 2010;116(2):451-8. Epub 2009/11/20. doi: 10.1002/cncr.24755. PubMed PMID: 19924789; PubMed Central PMCID: PMCPMC3188986.

22. Chaudhuri AA, Chabon JJ, Lovejoy AF, Newman AM, Stehr H, Azad TD, et al. Early Detection of Molecular Residual Disease in Localized Lung Cancer by Circulating Tumor DNA Profiling. Cancer Discov. 2017;7(12):1394-403. Epub 2017/09/14. doi: 10.1158/2159-8290.CD-17-0716. PubMed PMID: 28899864; PubMed Central PMCID: PMCPMC5895851.

23. Chin RI, Chen K, Usmani A, Chua C, Harris PK, Binkley MS, et al. Detection of Solid Tumor Molecular Residual Disease (MRD) Using Circulating Tumor DNA (ctDNA). Mol Diagn Ther. 2019;23(3):311-31. Epub 2019/04/04. doi: 10.1007/s40291-019-00390-5. PubMed PMID: 30941670; PubMed Central PMCID: PMCPMC6561896.

24. Corcoran RB, Chabner BA. Application of Cell-free DNA Analysis to Cancer Treatment. N Engl J Med. 2018;379(18):1754-65. Epub 2018/11/01. doi: 10.1056/NEJMra1706174. PubMed PMID: 30380390.

25. Wan JCM, Massie C, Garcia-Corbacho J, Mouliere F, Brenton JD, Caldas C, et al. Liquid biopsies come of age: towards implementation of circulating tumour DNA. Nat Rev Cancer. 2017;17(4):223-38. Epub 2017/02/25. doi: 10.1038/nrc.2017.7. PubMed PMID: 28233803.

26. Cristiano S, Leal A, Phallen J, Fiksel J, Adleff V, Bruhm DC, et al. Genome-wide cell-free DNA fragmentation in patients with cancer. Nature. 2019;570(7761):385-+. doi: 10.1038/s41586-019-1272-6. PubMed PMID: WOS:000472145700068.

27. Jiang P, Chan CW, Chan KC, Cheng SH, Wong J, Wong VW, et al. Lengthening and shortening of plasma DNA in hepatocellular carcinoma patients. Proc Natl Acad Sci U S A. 2015;112(11):E1317-25. Epub 2015/02/04. doi: 10.1073/pnas.1500076112. PubMed PMID: 25646427; PubMed Central PMCID: PMCPMC4372002.

28. Mouliere F, Chandrananda D, Piskorz AM, Moore EK, Morris J, Ahlborn LB, et al. Enhanced detection of circulating tumor DNA by fragment size analysis. Sci Transl Med. 2018;10(466). Epub 2018/11/09. doi: 10.1126/scitranslmed.aat4921. PubMed PMID: 30404863; PubMed Central PMCID: PMCPMC6483061.

29. Mouliere F, Mair R, Chandrananda D, Marass F, Smith CG, Su J, et al. Detection of cell-free DNA fragmentation and copy number alterations in cerebrospinal fluid from glioma patients. EMBO Mol Med. 2018;10(12). Epub 2018/11/08. doi: 10.15252/emmm.201809323. PubMed PMID: 30401727; PubMed Central PMCID: PMCPMC6284385.

30. Underhill HR, Kitzman JO, Hellwig S, Welker NC, Daza R, Baker DN, et al. Fragment Length of Circulating Tumor DNA. PLoS Genet. 2016;12(7):e1006162. Epub 2016/07/20. doi: 10.1371/journal.pgen.1006162. PubMed PMID: 27428049; PubMed Central PMCID: PMCPMC4948782.

31. Hirbe AC, Dahiya S, Miller CA, Li T, Fulton RS, Zhang X, et al. Whole Exome Sequencing Reveals the Order of Genetic Changes during Malignant Transformation and Metastasis in a Single Patient with NF1-plexiform Neurofibroma. Clin Cancer Res. 2015;21(18):4201-11. Epub 2015/05/01. doi: 10.1158/1078-0432.CCR-14-3049. PubMed PMID: 25925892; PubMed Central PMCID: PMCPMC4573781.

32. Pemov A, Hansen NF, Sindiri S, Patidar R, Higham CS, Dombi E, et al. Low mutation burden and frequent loss of CDKN2A/B and SMARCA2, but not PRC2, define pre-malignant neurofibromatosis type 1-associated atypical neurofibromas. Neuro Oncol. 2019. Epub 2019/02/06. doi: 10.1093/neuonc/noz028. PubMed PMID: 30722027; PubMed Central PMCID: PMCPMC6682216.

33. National Institutes of Health Consensus Development Conference Statement: neurofibromatosis. Bethesda, Md., USA, July 13-15, 1987. Neurofibromatosis. 1988;1(3):172-8. Epub 1988/01/01. PubMed PMID: 3152465.

34. von Elm E, Altman DG, Egger M, Pocock SJ, Gotzsche PC, Vandenbroucke JP, et al. The Strengthening the Reporting of Observational Studies in Epidemiology (STROBE) statement: guidelines for reporting observational studies. PLoS Med. 2007;4(10):e296. Epub 2007/10/19. doi: 10.1371/journal.pmed.0040296. PubMed PMID: 17941714; PubMed Central PMCID: PMCPMC2020495.

35. illumina. Mechanical DNA Fragmentation with the Q800R2 Sonicator: The Qsonica Q800R2 Sonicator offers an alternative method for mechanical DNA fragmentation for Illumina TruSeq Library Preparation Kits 2017 [cited 2020]. Available from: https://www.illumina.com/content/dam/illumina-marketing/documents/products/technotes/qsonica-instrument-tech-note-770-2016-030.pdf.

36. Pedersen BS, Quinlan AR. Mosdepth: quick coverage calculation for genomes and exomes. Bioinformatics. 2018;34(5):867–8. doi: 10.1093/bioinformatics/btx699. PubMed PMID: WOS:000426813500020.

37. Adalsteinsson VA, Ha G, Freeman SS, Choudhury AD, Stover DG, Parsons HA, et al. Scalable whole-exome sequencing of cell-free DNA reveals high concordance with metastatic tumors. Nat Commun. 2017;8(1):1324. Epub 2017/11/08. doi: 10.1038/s41467-017-00965-y. PubMed PMID: 29109393; PubMed Central PMCID: PMCPMC5673918.

38. Ramirez F, Ryan DP, Gruning B, Bhardwaj V, Kilpert F, Richter AS, et al. deepTools2: a next generation web server for deep-sequencing data analysis. Nucleic Acids Res. 2016;44(W1):W160-5. Epub 2016/04/16. doi: 10.1093/nar/gkw257. PubMed PMID: 27079975; PubMed Central PMCID: PMCPMC4987876.

39. Eisenhauer EA, Therasse P, Bogaerts J, Schwartz LH, Sargent D, Ford R, et al. New response evaluation criteria in solid tumours: revised RECIST guideline (version 1.1). Eur J Cancer. 2009;45(2):228-47. Epub 2008/12/23. doi: 10.1016/j.ejca.2008.10.026. PubMed PMID: 19097774.

40. Pemov A, Li H, Patidar R, Hansen NF, Sindiri S, Hartley SW, et al. The primacy of NF1 loss as the driver of tumorigenesis in neurofibromatosis type 1-associated plexiform neurofibromas. Oncogene. 2017;36(22):3168-77. Epub 2017/01/10. doi: 10.1038/onc.2016.464. PubMed PMID: 28068329; PubMed Central PMCID: PMCPMC6689395.

41. Upadhyaya M, Spurlock G, Thomas L, Thomas NS, Richards M, Mautner VF, et al. Microarray-based copy number analysis of neurofibromatosis type-1 (NF1)-associated malignant peripheral nerve sheath tumors reveals a role for Rho-GTPase pathway genes in NF1 tumorigenesis. Hum Mutat. 2012;33(4):763-76. Epub 2012/02/15. doi: 10.1002/humu.22044. PubMed PMID: 22331697.

42. Banerjee J, Allaway RJ, Taroni JN, Baker A, Zhang X, Moon CI, et al. Integrative Analysis Identifies Candidate Tumor Microenvironment and Intracellular Signaling Pathways that Define Tumor Heterogeneity in NF1. Genes (Basel). 2020;11(2). Epub 2020/02/27. doi: 10.3390/genes11020226. PubMed PMID: 32098059; PubMed Central PMCID: PMCPMC7073563.

43. Brohl AS, Kahen E, Yoder SJ, Teer JK, Reed DR. The genomic landscape of malignant peripheral nerve sheath tumors: diverse drivers of Ras pathway activation. Sci Rep. 2017;7(1):14992. Epub 2017/11/10. doi: 10.1038/s41598-017-15183-1. PubMed PMID: 29118384; PubMed Central PMCID: PMCPMC5678116.

44. Kryukov GV, Wilson FH, Ruth JR, Paulk J, Tsherniak A, Marlow SE, et al. MTAP deletion confers enhanced dependency on the PRMT5 arginine methyltransferase in cancer cells. Science. 2016;351(6278):1214–8. doi: 10.1126/science.aad5214. PubMed PMID: WOS:000386158400049.

45. Lee W, Teckie S, Wiesner T, Ran L, Prieto Granada CN, Lin M, et al. PRC2 is recurrently inactivated through EED or SUZ12 loss in malignant peripheral nerve sheath tumors. Nature Genetics. 2014;46(11):1227–32. doi: 10.1038/ng.3095.

46. Dehner C, Moon CI, Zhou Z, Zhang X, Miller C, Xu H, et al. Chromosome 8 gain is associated with high-grade transformation in MPNST. JCI Insight. 2021;6(6):e146351. doi: 10.1172/jci.insight.146351. PubMed Central PMCID: PMCPMC8026192.

47. Derlin T, Tornquist K, Munster S, Apostolova I, Hagel C, Friedrich RE, et al. Comparative effectiveness of 18F-FDG PET/CT versus whole-body MRI for detection of malignant peripheral nerve sheath tumors in neurofibromatosis type 1. Clin Nucl Med. 2013;38(1):e19-25. Epub 2012/12/18. doi: 10.1097/RLU.0b013e318266ce84. PubMed PMID: 23242059.

48. Ferner RE, Gutmann DH. International consensus statement on malignant peripheral nerve sheath tumors in neurofibromatosis. Cancer Res. 2002;62(5):1573-7. Epub 2002/03/16. PubMed PMID: 11894862.

49. Reilly KM, Kim A, Blakely J, Ferner RE, Gutmann DH, Legius E, et al. Neurofibromatosis Type 1-Associated MPNST State of the Science: Outlining a Research Agenda for the Future. J Natl Cancer Inst. 2017;109(8). Epub 2017/11/09. doi: 10.1093/jnci/djx124. PubMed PMID: 29117388; PubMed Central PMCID: PMCPMC6057517.

50. Akshintala S, Baldwin A, Liewehr DJ, Goodwin A, Blakeley JO, Gross AM, et al. Longitudinal evaluation of peripheral nerve sheath tumors in neurofibromatosis type 1: growth analysis of plexiform neurofibromas and distinct nodular lesions. Neuro Oncol. 2020;22(9):1368-78. Epub 2020/03/11. doi: 10.1093/neuonc/noaa053. PubMed PMID: 32152628; PubMed Central PMCID: PMCPMC7523449.

51. Canavese F, Krajbich JI. Resection of plexiform neurofibromas in children with neurofibromatosis type 1. J Pediatr Orthop. 2011;31(3):303-11. Epub 2011/03/19. doi: 10.1097/BPO.0b013e31820cad77. PubMed PMID: 21415691.

52. Mautner VF, Hartmann M, Kluwe L, Friedrich RE, Funsterer C. MRI growth patterns of plexiform neurofibromas in patients with neurofibromatosis type 1. Neuroradiology. 2006;48(3):160-5. Epub 2006/01/25. doi: 10.1007/s00234-005-0033-4. PubMed PMID: 16432718.

53. Wasa J, Nishida Y, Tsukushi S, Shido Y, Sugiura H, Nakashima H, et al. MRI Features in the Differentiation of Malignant Peripheral Nerve Sheath Tumors and Neurofibromas. Am J Roentgenol. 2010;194(6):1568–74. doi: 10.2214/Ajr.09.2724. PubMed PMID: WOS:000277948400021.

54. Chabon JJ, Hamilton EG, Kurtz DM, Esfahani MS, Moding EJ, Stehr H, et al. Integrating genomic features for non-invasive early lung cancer detection. Nature. 2020;580(7802):245-51. Epub 2020/04/10. doi: 10.1038/s41586-020-2140-0. PubMed PMID: 32269342.

55. Cohen JD, Li L, Wang Y, Thoburn C, Afsari B, Danilova L, et al. Detection and localization of surgically resectable cancers with a multi-analyte blood test. Science. 2018;359(6378):926-30. Epub 2018/01/20. doi: 10.1126/science.aar3247. PubMed PMID: 29348365; PubMed Central PMCID: PMCPMC6080308.

56. Liu MC, Oxnard GR, Klein EA, Swanton C, Seiden MV, Consortium C. Sensitive and specific multi-cancer detection and localization using methylation signatures in cell-free DNA. Ann Oncol. 2020;31(6):745-59. Epub 2021/01/29. doi: 10.1016/j.annonc.2020.02.011. PubMed PMID: 33506766.

57. Shen SY, Singhania R, Fehringer G, Chakravarthy A, Roehrl MHA, Chadwick D, et al. Sensitive tumour detection and classification using plasma cell-free DNA methylomes. Nature. 2018;563(7732):579-+. doi: 10.1038/s41586-018-0703-0. PubMed PMID: WOS:000450960000058.

58. Chen X, Chang CW, Spoerke JM, Yoh KE, Kapoor V, Baudo C, et al. Low-pass Whole-genome Sequencing of Circulating Cell-free DNA Demonstrates Dynamic Changes in Genomic Copy Number in a Squamous Lung Cancer Clinical Cohort. Clin Cancer Res. 2019;25(7):2254-63. Epub 2019/01/09. doi: 10.1158/1078-0432.CCR-18-1593. PubMed PMID: 30617129.

